# Early transcriptional changes in neutrophil-mediated processes following recanalization after ischemic stroke

**DOI:** 10.1101/2024.11.05.24316798

**Authors:** Truong An Bui, Yonglie Ma, Glen C Jickling, Ian R Winship

## Abstract

**Background:** Ischemic stroke is a leading cause of mortality and long-term disability globally. Recanalization therapies restore blood flow by thrombolysis and/or mechanical thrombectomy. Despite successful recanalization, many patients still experience poor clinical outcomes. This phenomenon, known as futile recanalization, may result from reperfusion injury and microcirculatory failure. Given the short therapeutic window for ischemic stroke, it is critical to identify early biomarkers that could be targeted to minimize pathology and extend the therapeutic window to improve clinical outcomes.

**Methods:** Using a murine middle cerebral occlusion (MCAO) model that mimics a large vessel occlusion with recanalization, a comprehensive microarray analysis of gene expression from blood samples collected immediately and 3 hours after recanalization (N=44) was performed. Differentially expressed genes (DEGs), enrichment pathways, immune cell proportions, enriched cell markers, and predicted miRNAs and transcription factors were identified using RStudio. Findings in mice were validated in datasets of rat MCAO stroke (GSE21136) and human stroke patients (GSE16561) to confirm transcriptional changes in peripheral blood post-recanalization.

**Results:** *Il1r2, Cd55, Mmp8, Cd14,* and *Cd69* were identified as early biomarkers in blood after stroke and recanalization. Further, cross-validation revealed *Vcan* as a DEG conserved across species, making it a novel marker of ischemia detected as early as 3 hours post-recanalization (4 hours post-MCAO) in mice, 24 hours after recanalization in rats (MCAO-thrombectomy), and within 24 hours from onset in humans receiving rtPA-thrombolysis. Using human (CIBERSORTx) and murine (ImmuCellAI-mouse) cell deconvolution reference datasets, neutrophil was elevated post-recanalization. Leukocyte and neutrophil activation pathways were significantly enriched early after stroke in mouse and human samples, with a stronger upregulation observed in females of both species. The analysis revealed several key miRNAs involved in gene regulation following recanalization; NFE4 and MTF1 emerged as essential transcription factors in these processes. Based on these data, a coregulatory network underlying neutrophil activity was constructed, highlighting its central role in early responses to ischemia and recanalization, which was enriched in females.

**Conclusions:** This study identified new early genomic markers in the blood for ischemia and recanalization, as well as critical age- and sex-specific factors. By mapping a coregulatory network of interacting genes and neutrophil-related pathways, as well as identifying the novel marker *Vcan*, the data provides insights to inform future research and develop targeted therapies. Such therapies can improve treatment efficacy or modulate neutrophils to reduce futile recanalization, ultimately enhancing clinical outcomes for ischemic stroke patients.

**Graphical Abstract:** 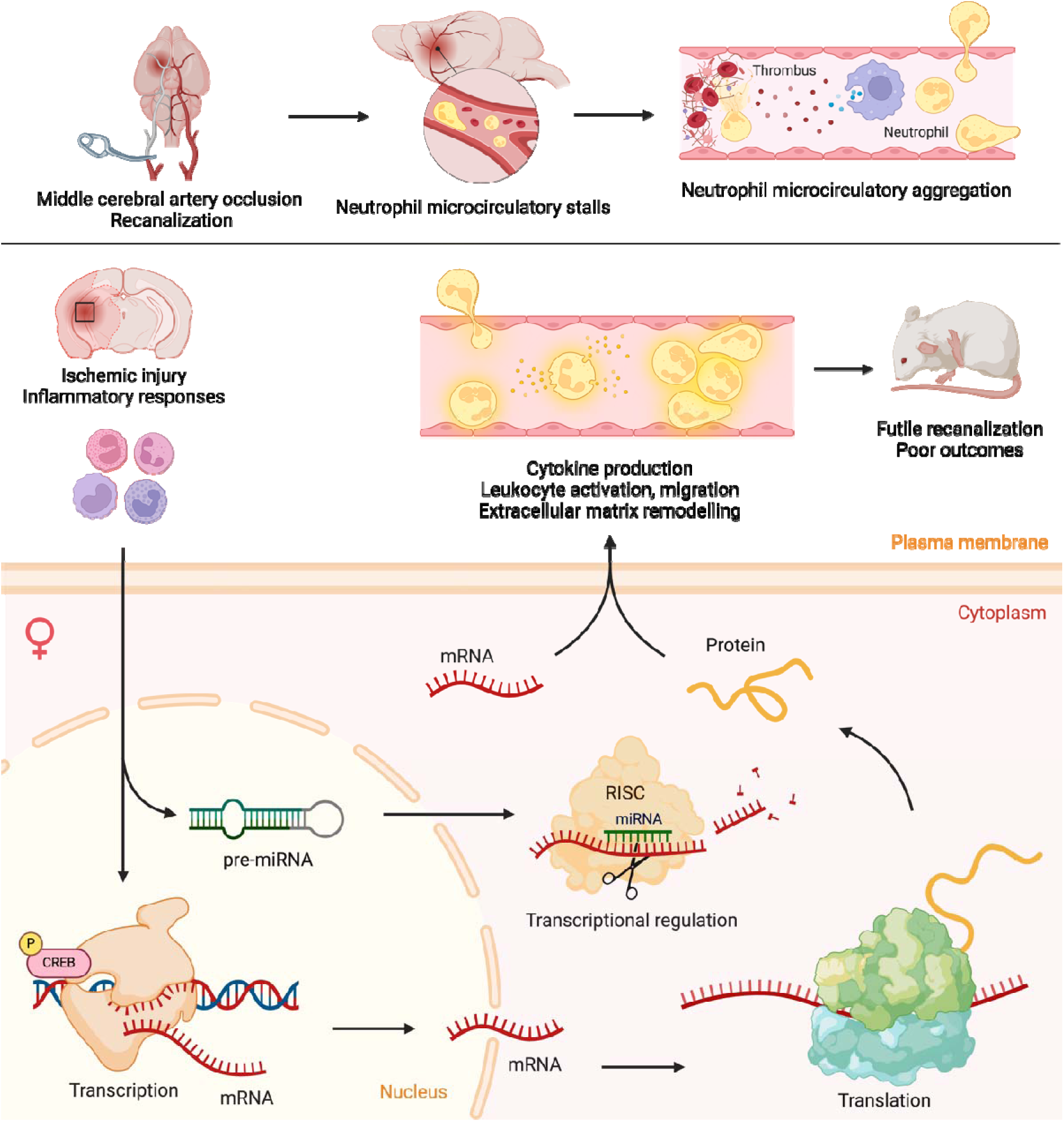

**Clinical and research perspectives:** This study identifies *Il1r2, Cd55, Mmp8, Cd14, Cd69*, and *Vcan* as novel, early blood biomarkers of ischemic stroke and reperfusion injury following recanalization. Notably, *Vcan* was conserved across rodent and human models, underscoring its translational potential. These peripheral hemo-genomic signatures hold promise as biomarkers to help identify patients at higher risk of futile recanalization, thereby enabling earlier, more tailored clinical interventions to reduce reperfusion injury and improve recovery. The significant increase in neutrophil proportions, demonstrated through deconvolution of both human and murine reference datasets, supports prior evidence implicating neutrophil aggregation in reperfusion failure and highlights this cell type as a key target for further investigation. Moreover, the pronounced neutrophil-related transcriptional responses in females suggest that sex-specific mechanisms may influence stroke outcomes and warrant consideration in therapeutic design in acute stroke management. At the mechanistic level, the study reveals a core gene regulatory network—centred on neutrophil-associated genes, transcription factors NFE4 and MTF1, and stroke-associated miRNAs—that orchestrates early immune responses to ischemia and recanalization. Collectively, these findings suggest potentially predictive molecular signatures of reperfusion failure following recanalization. Future studies could investigate whether targeting these regulatory elements can prevent futile recanalization, thus improving long-term functional outcomes.

## Introduction

In the 2021 Global Burden of Diseases, Injuries, and Risk Factors Study, stroke was identified as the third leading cause of mortality worldwide, following ischemic heart disease and COVID-19 (1). Ischemic stroke, the predominant subtype, accounts for approximately 75-80% of all stroke cases (2). In 2021, stroke also resulted in an estimated 3,870,000 deaths globally (1,3) and contributed to 160.4 million disability-adjusted life years (4). The epidemiology of stroke reveals a pronounced age-related incidence, predominantly affecting individuals aged 65 and older (2). Notably, there is a sex disparity in the age of onset, with women experiencing strokes 4 to 6 years later than men (5). However, women under 35 years old have a 44% higher likelihood of ischemic stroke relative to men and women overall have a greater lifetime risk of ischemic events and transient ischemic attacks (6). After middle age, the incidence and long-term risks of ischemic stroke rise more steeply in women, paralleled by an increase in the severity of resultant disabilities (7). This heightened risk with increasing age contributes to a significant public health challenge, given the global demographic shifts towards the aging populations (8).

The best available treatment for ischemic stroke is recanalization therapy via thrombolysis with recombinant tissue plasminogen activator (alteplase) (r-tPA) or tenecteplase (9) and/or endovascular therapy (EVT) (10). While recanalization restores flow in the occluded vessel, this does not always extend into the microcirculation that nourishes brain tissue. This can result from collateral failure, whereby cerebral ischemia in penumbral regions worsens over time due to the collapse of the collateral circulation (11). It also results from microcirculatory failure, where dysfunction in the small blood vessels within the brain tissue impairs microcirculation, despite the large vessels being recanalized (12). A key contributor to this dysfunction is the post-ischemic “stalling” of neutrophils within cerebral microvessels. After recanalization, neutrophils can adhere to endothelial cells and aggregate in microvessels, thereby preventing blood flow, becoming activated, and releasing pro-inflammatory cytokines, which amplify tissue damage and drive further inflammation (for review, see (13)).

Ischemic stroke progression follows a well-defined time course, beginning with the immediate occlusion of a major cerebral artery, leading to a rapid depletion of oxygen and glucose, energy failure, and excitotoxicity in the infarct core, while the surrounding penumbra remains metabolically active but vulnerable (14–16). Without timely intervention, the penumbra undergoes irreversible damage as collateral circulation fails and inflammation escalates (17). The therapeutic window for recanalization is highly restricted—thrombolysis with r-tPA is only approved within 4.5 hours of stroke onset, and EVT is most effective within 6-24 hours for select patients (18–20). Furthermore, while recanalization aims to restore blood flow, alleviate tissue hypoxia, and salvage ischemic tissue, restoration of flow can paradoxically cause tissue damage. This complication, known as reperfusion injury, happens when blood flow is abruptly restored to previously ischemic tissue after a period of oxygen deprivation. This causes a rush of molecular oxygen, leading to the rapid production of reactive oxygen species. This, in turn, causes oxidative stress, prolonged inflammation, and activation of immune cells in the blood and brain and can potentiate microcirculatory failure (21).

Although the success rate of recanalization has improved over time, many patients still experience poor long-term outcomes, as measured by the modified Rankin Scale (mRS) at 90 days and the National Institutes of Health Stroke Scale. This phenomenon, known as futile recanalization—where patients remain functionally dependent despite successful recanalization (mRS ≥ 3)—occurs in 32-57% of ischemic stroke patients who undergo EVT (22,23) and is exacerbated by collateral failure, microcirculatory failure, and reperfusion injury (11,24). Patterns of gene expression following ischemic stroke offer valuable insights into the underlying causes of reperfusion injury and potential outcomes of recanalization. Comparative analyses of gene expression profiles across different types of blood cells in stroke patients reveal that transcriptional alterations primarily occur in neutrophils as early as 15 minutes after ischemic onset (25). These transcriptional shifts are modulated by interactions among neutrophils, platelets, and cytokines, which can amplify the inflammatory response and influence stroke outcomes (13). This suggests neutrophils can be an early peripheral and genomic signature of post-recanalization microcirculatory failure (26). Changes in gene expression profiles may indicate ongoing inflammatory and repair processes, potentially serving as biomarkers to assess the severity and progression of stroke, as well as the risks and mechanisms of futile recanalization.

There remains a critical need to identify novel biomarkers that can reliably predict which patients are at the highest risk of microcirculatory failure and futile recanalization. Current clinical assessment tools are insufficient in capturing the complex interplay between immune responses, endothelial dysfunction, and reperfusion injury at a molecular level (27). A deeper understanding of the transcriptomic landscape of circulating immune cells, particularly neutrophils, could reveal novel prognostic markers and therapeutic targets to mitigate post-recanalization complications. Moreover, integrating high-throughput gene expression analyses with single-cell RNA sequencing and proteomics could uncover previously unrecognized pathways that drive neutrophil-mediated microvascular dysfunction.

A rapidly measurable blood-based biomarker panel is invaluable for informing clinical decisions during initial triage and acute care to reduce the risks of futile recanalization. By incorporating novel molecular markers into point-of-care diagnostics, clinicians can better stratify patients based on their likelihood of responding to recanalization therapies and tailor interventions accordingly. Future studies should focus on identifying gene expression patterns that differentiate beneficial immune responses from pathological inflammation, paving the way for targeted therapies that could extend the therapeutic window and improve stroke recovery outcomes.

This study explored early transcriptional changes occurring in peripheral blood following stroke with or without recanalization in a mouse model. Dynamic gene expression patterns were analyzed to uncover regulatory pathways responding to MCAO and recanalization. The analyses identified several critical genes that could serve as new biomarkers or predictors of ischemic/reperfusion injury following recanalization. Notably, sex- and age-specific differences in gene expression were observed, with female mice and aged mice displaying unique leukocyte and neutrophil-related gene activation patterns. *Vcan* emerged as a novel biomarker for ischemic stroke and recanalization, and the only gene conserved across mice, rats, and humans. *Vcan*’s distinct expression in aged animals, especially as a central miRNA regulatory node, points to a previously unexplored aspect of ischemic stroke and reperfusion injury biology. Finally, a coregulatory network composed of rich interactions between identified differentially expressed genes, miRNAs, and their transcription factors in neutrophil-centred pathways linked to reperfusion injury was compiled for the first time. These discoveries open new avenues for understanding the molecular mechanisms driving stroke recovery and may provide insights for developing therapies tailored to age and sex.

## Methods

### 1. Middle cerebral artery occlusion (MCAO) surgery

Experimental methods are outlined in **Supplementary Figure 1**. Wild-type mice (C57BL/6J) were obtained from the University of Alberta North Campus Animal Services and housed in the university facility’s pathogen-free animal room at 18-22°C and 60% humidity, with free access to food and water. Standard diet and husbandry conditions were maintained. Proximal middle cerebral artery occlusion (MCAO) was modelled by transecting the external carotid artery, ligating the common carotid artery, and inserting a Doccol monofilament through the internal carotid artery to occlude the origin of the middle cerebral artery. The MCAO model closely mimics the pathophysiology of human ischemic stroke, blocking the major middle cerebral artery, with controlled recanalization/reperfusion. Body temperature was monitored throughout the surgery using a rectal probe and maintained at 37.0 ± 0.5°C by a heating pad. The suture was placed for 60 minutes before removal to mimic complete recanalization.

Three experimental groups (N=44), sham (SHAM, N=16), 1-hour MCAO (MCAO, N=17), and 1-hour MCAO with 3-hour recovery post-recanalization (MCAO+Recan, N=11), were included in the dataset. Both young adult (3-6 months old, N=20) and aged (18-24 months old, N=24), male (N=20) and female (N=24) mice were used. Randomization was used to assign treatment. To limit potential confounders, each cage also has all SHAM, MCAO, and MCAO+Recan groups. MCAO mice were euthanized immediately 60 minutes after surgery, blood samples were collected, and the suture causing MCAO was removed. In the MCAO+Recan group, the suture was removed 60 minutes after the onset of occlusion and mice were allowed to recover for an additional 3 hours before euthanasia. SHAM mice were subjected to only anesthesia and the manipulation of surrounding tissues and arteries, with 4 hours of recovery before sample collection. Saline and buprenorphine (0.03mg/kg) were administered subcutaneously after MCAO surgery for pain management. Animals were kept under anesthesia during surgery (2.0-2.5% isoflurane (70:30, O_2_:N_2_O)).

All mice were anesthetized with an intraperitoneal injection of 20% urethane at the point of sample collection. Blood samples were obtained immediately with retro-orbital sampling, collected in 500µL RNAprotect animal blood tubes (Qiagen, Germany), and stored at −20°C until RNA purification. Mice were then euthanized by decapitation. ARRIVE (Animal pre-clinical studies) checklist was completed (**Supplementary Table 3**) (28). All experiments were approved by the University of Alberta’s Health Sciences Animal Care and Use Committee (Protocol AUP361) and adhered to the guidelines set by the Canadian Council for Animal Care.

### 2. Sample collection and microarray analysis

Total RNA was isolated from blood samples using the RNeasy Kits, a process that was automated by the QIAcube (Qiagen, Germany). RNA was cleaned, and contaminants were removed using the RNeasy MinElute CleanUp Kit (Qiagen, Germany). NanoDrop 2000 (Thermo Fisher Scientific, U.S.) and Agilent Bioanalyzer 2100 (Agilent Technologies, U.S.) were used to measure RNA yield and detect integrity before microarray analysis. Samples with an RNA integrity number (RIN) of less than 7, A260/A280 ratio of less than 1.8, and volume of less than 35uL each at 50ng/uL were excluded from the dataset (ThermoFisher Scientific, U.S.). Reads were measured by Affymetrix Clariom S microarray (Affymetrix, ThermoFisher Scientific) by North American Genomics. Experimenters and technicians were blinded during sample processing. 58 blood samples were obtained, and 44 samples passed RIN, A260/A280 ratio, and volume cutoff.

### 3. Expression data analysis

All expression analysis was performed in RStudio. The limma package, which allows for variance stabilization, was used to normalize reads and identify differentially expressed genes (DEGs) across sex and age from raw data CEL files (|log_2_FC| ≥1.5, p-vale ≤ 0.05) (29). An ExpressionSet object containing expression and phenotype data was created and used in all downstream analyses. Exploratory data analysis techniques, including heatmaps, principal component analysis (PCA), and clustering, were employed to examine data. The volcano plot and heatmap for DEGs were plotted with the R packages ggplot2 and ComplexHeatmap. Sample clustering was performed. We used Pearson’s Chi-squared test with Yates’ continuity correction to evaluate the associations between these sample clusters and the categorical variables of treatment, sex, and age.

DEGs were exported to Cytoscape Version 3.9.1 to visualize their interactive network. Protein-protein interaction networks (PPIs) of DEGs were constructed with STRING (https://string-db.org). MCODE V1.5.1 was used to identify significant modules (MCODE score ≥ 4). Hub genes were selected using CytoHubba. GeneCards was used to look up previous finding and gene functions (30). R/Bioconductor package gProfiler was used for functional enrichment analysis. gProfiler computes the enrichment of a pathway from the input gene list using Fisher’s exact test and multiple-test correction (31). NetworkAnalyst platform and miRTarbase were used to predict miRNA–gene interaction (32). ChIP Enrichment Analysis (ChEA) was used to obtain transcription factors (TFs). ChEA results help identify candidate TFs that likely regulate the gene set based on experimentally derived TF target profiles, along with statistics and gene overlap details, but without TF expression, activity status, or mechanisms (33). Additional graphs were visualized in BioRender.

### 4. Integrative cellular profiling

Both deconvolution methods (CIBERSORT for humans and ImmuCellAI for mice) and cell marker dataset enrichment (CellMarker) were used since they provide complementary insights into the cellular composition and transcriptional activity of bulk microarray samples.

Cell Marker 2.0 and clusterProfiler were used to infer the cell types in which the DEGs might be found (34,35). Cell-type associations were identified based on lists of DEGs. Upregulated and downregulated DEGs for each condition were separately analyzed using over-representation analysis (ORA) via enricher, mapping to annotated cell types. This method enables a deeper understanding of the functional changes within specific cell types or to identify potential cell types that contribute to differential gene expression.

To estimate the relative abundance of known immune cell populations in our samples, we employed both ImmuCellAI-mouse (36) and CIBERSORTx using the LM22 leukocyte gene signature matrix (37–40). ImmuCellAI-mouse was selected for its optimized performance in quantifying 36 distinct mouse immune cell subtypes, providing high-resolution deconvolution tailored to our murine datasets (36). In parallel, CIBERSORTx, which leverages the human-specific LM22 signature, was applied to facilitate cross-species comparison through orthologous gene mapping (37–40). Analyses were conducted in high-resolution mode, restricted to protein-coding transcripts to enhance specificity. The resulting immune cell proportion estimates from both platforms were statistically compared across experimental groups using the Kruskal–Wallis test with Benjamini-Hochberg adjustment, followed by Dunn’s post hoc correction, with a false discovery rate (FDR) threshold of 0.05 (41,41,42). Aligned rank transform analysis of variance (ART ANOVA) with Benjamini-Hochberg adjustment (FDR ≤ 0.05) was used to identify interactions between treatment, species differences, age, and sex that influence changes in cell proportions. These combined strategies obtain a comprehensive picture: quantitative cell type proportions from deconvolution, and mechanistic insights into cell type-specific transcriptional activity from marker enrichment, ultimately strengthening the ability to interpret complex biological differences between mouse and human models.

### 5. Cross-validation

The findings were compared to those from previous studies in the Gene Expression Omnibus (GEO) database to validate the results. The search query [ischemic stroke AND (reperfusion OR middle cerebral occlusion) AND peripheral blood] was used to obtain studies surveying transcriptional changes in peripheral blood post-recanalization. Datasets GSE16561 (43) and GSE21136 (44) were collected from the GEO database.

GSE16561 comprises clinical data analyzing blood expression from patients with MRI-diagnosed ischemic stroke, including those who received rt-PA (within 3 hours from onset) and controls (43). The median time from symptom onset to blood draw in these subjects was 7.4 hours (45). GSE21136 includes microarray analysis on rats that received MCAO+Recan with and without rtPA (44). Similar differential gene expression and functional enrichment analyses were performed on these datasets in R. All data in this study are available in the GEO database under accession number GSE278554.

## Results

Following the reading of raw CEL files and normalization with rma, the entire dataset was assigned to an ExpressionSet object, which consists of 44 samples and 29,129 quantified transcripts. A heatmap plot identified four clusters of genes, with clusters seeming to correspond primarily to sex differences **(Figure 1A**). Pearson’s Chi-squared test with Yates’ continuity correction was conducted to determine whether sample clustering based on gene expression profiles was significantly associated with treatment, sex, or age. Treatment (χ² = 2.7904, df = 2, p-value = 0.2478), age (χ² = 0.23653, df = 1, p-value = 0.6267), and sex (χ² = 1.2449, df = 1, p-value = 0.2645), however, were not significantly associated with cluster sampling.

**Figure 1.**
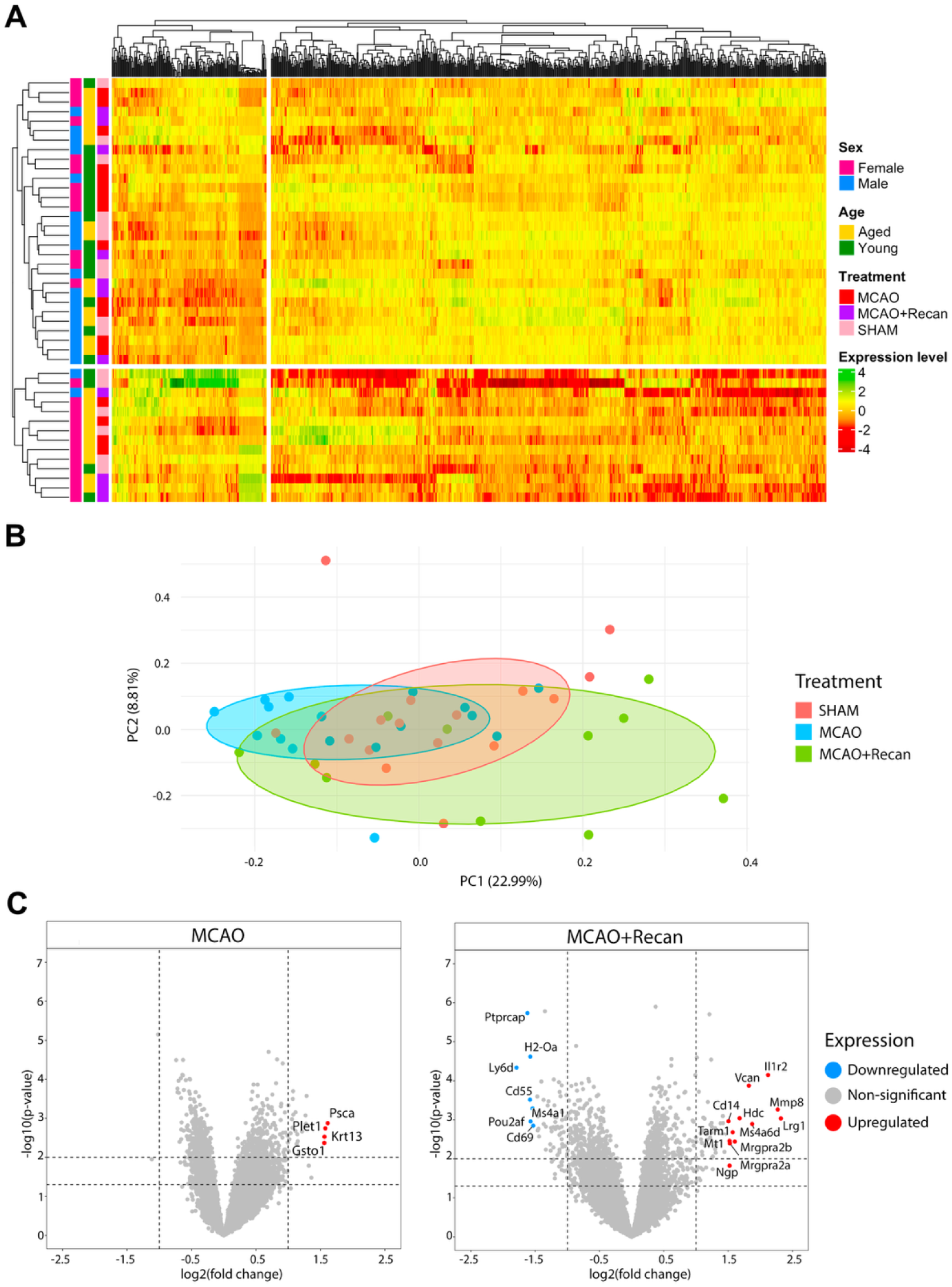
**(A)** Hierarchical clustering of gene expression patterns across all samples. The cluster dendrograms represent Canberra distance-based hierarchical clustering, which organizes samples (columns) and genes (rows) based on similarities in their expression. The dendrograms on the left and top of the heatmap display the clustering structure, visually grouping samples and genes with similar expression profiles. **(B)** Principal component analysis (PCA) of 44 samples. The PCA plot illustrates the expression variability among the samples, with each point representing the combined expression of all differentially expressed genes (DEGs) for a given sample. Samples are grouped by experimental condition (SHAM, MCAO, and MCAO+Recan), as indicated by the coloured ellipses. The x-axis represents the first principal component, which explains 22.99% of the total variance, while the y-axis represents the second principal component, explaining 8.81% of the variance. **(C)** Volcano plots of DEGs in MCAO and MCAO+Recan group. The plots depict the significantly upregulated (red) and downregulated (blue) genes in the MCAO (left) and MCAO+Recan (right) groups (|log2FC| ≥ 1.5, p-value ≤ 0.05). The plots compare the magnitude and significance of gene expression changes. Differential gene expression analysis was performed using the limma package in RStudio and Bioconductor.

PCA showed that the SHAM group and the MCAO group (i.e. MCAO with blood sampled 1 hour after ischemic onset, prior to recanalization) were closely clustered and had less internal variation. In contrast, the MCAO+Recan group (i.e. blood samples 3 hours after recanalization or 4 hours after ischemic onset) has a greater internal variation and heterogeneous responses (**Figure 1B**). Nevertheless, PC1 (22.99%) and PC2 (8.81%) indicate that these biological variations were not significant.

Using an exploratory approach to allow for a broader survey of potential associations or relationships in these early transcriptional changes, differential gene expression (MCAO vs. SHAM and MCAO+Recan vs. SHAM) was analyzed. The comparison MCAO+Recan vs. MCAO (Recan group) was also examined for any notable recanalization effect. DEGs with a less conservative filter (|log_2_FC| ≥ 1.5, p ≤ 0.05) were identified and reported with adjusted p-values along with raw p-values (**Supplementary Table 1**).

### 1. Differential expression analysis

#### i. Identification of novel early ischemic stroke and recanalization markers

Differential gene expression analysis identified 4 significantly upregulated genes in the MCAO group compared to SHAM: *Plet1, Krt13, Psca,* and *Gsto1* (|log_2_FC| ≥ 1.5, p ≤ 0.05) (**Figure 1C**; **Supplementary Table 1**). These genes have been previously associated with tumour, cancer, and tissue damage (46–49). *Plet1* is a progenitor cell marker predicted to be involved in cell-matrix adhesion, cell migration, and wound healing (47). *Krt13* is related to inflammation, tissue damage, and epithelial proliferation and differentiation (50). *Gsto1* has previously been associated with inflammatory responses (51). However, to date, these genes, which showed significant upregulation only 1 hour after ischemic onset, have not been associated with ischemic stroke.

Samples from the MCAO+Recan group had an increased number of DEGs, with 12 significantly upregulated genes (*Il1r2, Hdc, Tarm1, Mrgpra2a, Mrgpra2b, Mt1, Mmp8, Ngp, Vcan, Lrg1, Cd14,* and *Ms4a6d*) and 7 significantly downregulated genes (*Cd55, Cd69, Pou2af1, Ly6d, H2-Oa, Ptprcap*, and *Ms4a1*) (|log_2_FC| ≥ 1.5, p ≤ 0.05) (**Figure 1C**; **Supplementary Table 1**). Among these, *Il1r2* (52,53)*, Hdc* (54,55)*, Mt1* (56)*, Mmp8* (57–59)*, Ngp* (60–62)*, Vcan* (63–65)*, Lrg1* (66–68)*, Cd14* (69–71)*, Ms4a4a* (72–74)*, Ms4a6d* (75–77), and *Cd69* (78–80) have been previously linked to ischemic stroke and inferred in immune-related pathogenesis but have not been identified as early as 4 hours after ischemic onset or after recanalization.

Additionally, Recan group (MCAO+Recan vs. MCAO) had 19 DEGs: *B3gnt5, Ms4a1, Slc15a4, Blk, Stap1, Nfkbid, Srpk1, Cd55, Ly6d, P2ry10b, Pgap1, Cd69, Pou2af1, Cd79b, Vcan, P2ry10, Ifi203, Lnx2,* and *Ms4a6d*, 7 of which were shared with MCAO+Recan group (**Supplementary Figure 2A**). The majority of DEGs in Recan were downregulated, especially B-cell and immune signalling genes, such as *Ms4a1, Cd79b, Pou2af1, Cd55, Cd69*, and *Blk*. *Vcan* and *Ms4a6d* were upregulated, implicating ECM changes and myeloid responses (**Supplementary Table 1**). *Vcan* is consistently upregulated in both MCAO+Recan and Recan groups, supporting its role in tissue remodelling and neuroinflammation post-stroke and recanalization. *Cd55* and *Cd69* were downregulated in both MCAO+Recan and Recan, indicating reduced T-cell and complement activity with reperfusion. *Ms4a1* (B-cell marker) was significantly downregulated in both MCAO+Recan and Recan, suggesting reperfusion might suppress adaptive immune responses (**Supplementary Table 1**).

We also performed multivariate analyses to examine the two-way interactions of Sex × Treatment and Age × Treatment. There were no significant interactions in the MCAO group; however, in the MCAO+Recan, we found 127 DEGs showing Age × Treatment interactions and 14 DEGs showing Sex × Treatment (|log_2_FC| ≥ 1.5, FDR ≤ 0.05) (**Supplementary Table 1**). These two groups share 9 DEGs: *Nexmif, Or8d23, Fam171b, Map3k1, Klc1, Vmn1r70, Dsg1b, Atp8b5,* and *Dgkz.* Only *Mrgpra2a* was shared between MCAO+Recan and Sex × MCAO+Recan with FDR < 0.05 cutoff. When using p-value < 0.05, Sex × MCAO+Recan shared *Mrgpra2a, Mrgpra2b, Lrg1, Tarm1, Mmp8, Mt1, Hdc,* and *Pou2af1* with MCAO+Recan and Age × MCAO+Recan shared *Cd14* with MCAO+Recan (**Supplementary Figure 2B**)

#### ii. Females and young animals showed greater transcriptional changes

We next examined how sex and age influence transcriptional changes following ischemia. Sex differences were evident. In the MCAO group, females had more DEGs than males (31 vs. 0, the latter had 12 differentially expressed transcripts with no mapped genes). In the MCAO+Recan group, females exhibited 118 DEGs, while males had only 9. Females in the MCAO and MCAO+Recan groups shared 14 DEGs, including *Asprv1, Cbr2, Cxcr2, Emp1, Gsto1, Hdc, Il1b, Il1rn, Lrg1, Mmp8, Mrgpra2a, Plet1, Prg4*, and *Sirpb1a*. Additionally, the female MCAO+Recan group shared *Ly6d* and *Ptprcap* with their male counterparts (**Figure 2A**).

**Figure 2.**
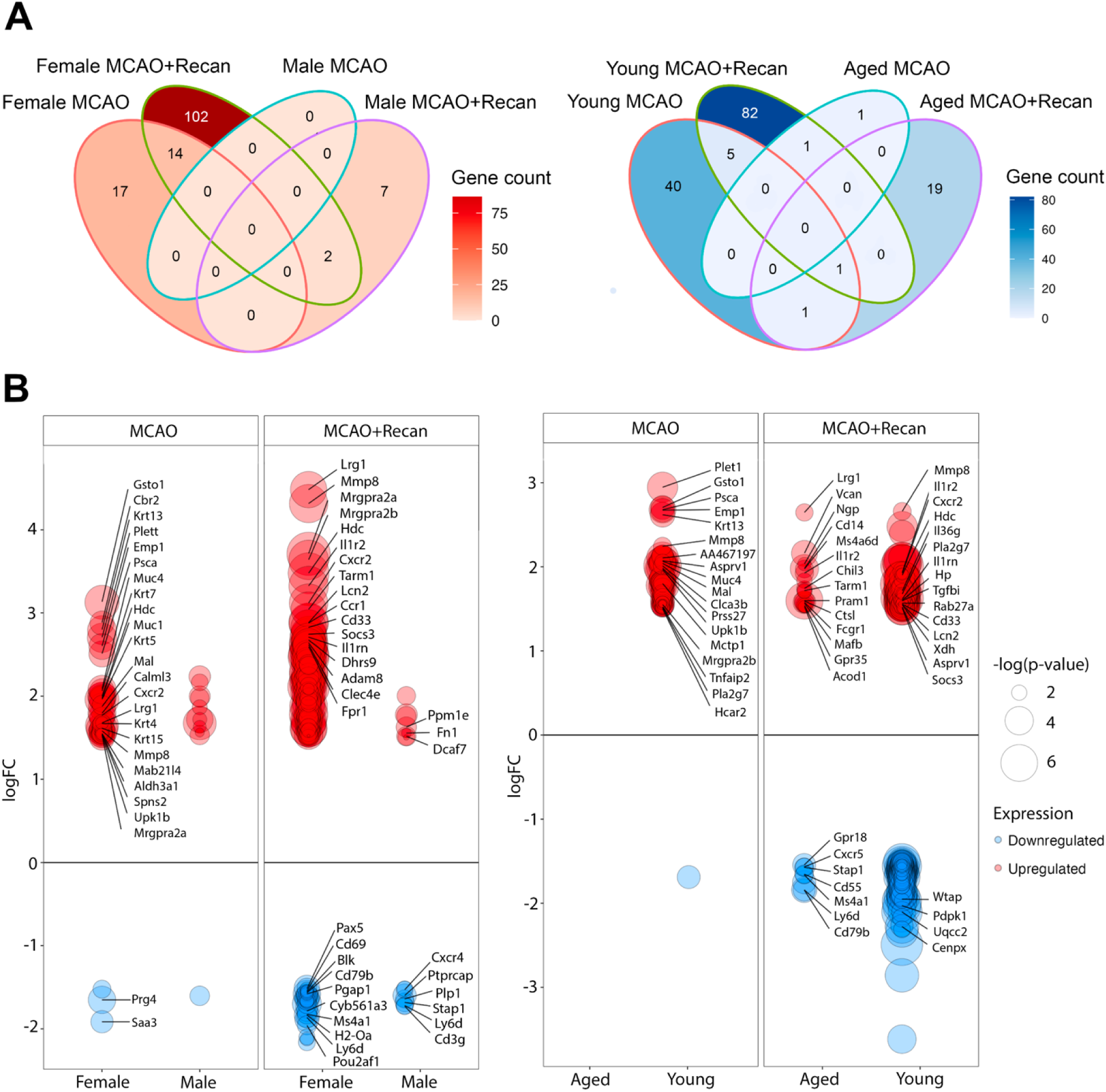
Age- and sex-specific differentially expressed genes (DEGs). Differential gene expression analysis was performed using the limma package from RStudio and Bioconductor. **(A)** Venn diagram showing the overlap of DEGs between sex-specific and age-specific groups (|log_2_FC| ≥ 1.5, p-value ≤ 0.05). **(B)** Bar plots depict the up and downregulated genes in age-specific and sex-specific groups (|log_2_FC| ≥ 1.5, p-value ≤ 0.05). These analyses highlight the unique and shared transcriptional changes based on age and sex in MCAO+Recan models.

This trend persisted when comparing MCAO+Recan with Recan groups: female Recan animals had 85 DEGs, 44 of which overlapped with female MCAO+Recan DEGs: *Mrgpra2a, Retnlg, Lcn2, Mrgpra2b, Il1r2, Ccr1, Mmp8, Il13ra1, Lrg1, Fpr2, Socs3, Cd33, Tarm1, Ifitm6, Bst1, Clec4e, Il1rap, Vcan, Nfyb, S100a8, Ptprcap, Cd14, Trim7, Ly6d, Cd83, Cyb561a3, Chil3, Ms4a1, Ms4a6d, Pou2af1, Cd55, Ngp, Nop56, Wfdc21, Mafb, Ets1, Mt1, Cxcr5, Macf1, Pax5, Cd79b, Pgap1, Blk*, and *Cd69*. On the contrary, male MCAO+Recan and Recan only shared 4 DEGs, including *Stap1, Plp1, Cd3g,* and *Cxcr4*. Between female and male Recan groups, 5 DEGs overlapped: *Ms4a1, B3gnt5, Srpk1, Pgap1,* and *Cd69*. Across all conditions, MCAO+Recan animals consistently showed the highest number of DEGs and females had more extensive transcriptional changes compared to males (**Supplementary Figure 2C**).

Overall, young animals displayed more pronounced transcriptional responses than aged animals. In the MCAO group, young animals had 47 DEGs compared to only 2 in aged animals. Similarly, in the MCAO+Recan group, young animals exhibited 89 DEGs, whereas aged animals had 21. Thus, at both 1 and 4 hours after ischemic onset, young animals showed substantially stronger gene expression changes than aged animals. When comparing MCAO and MCAO+Recan groups, young animals shared 6 common DEGs: *Asprv1, Hdc, Il1r2, Il1rn, Mmp8,* and *Pla2g7*; whereas aged animals had no common genes. When comparing young and aged animals in the MCAO+Recan group, the only shared gene is interleukin-1 receptor type II (*Il1r2*) (**Figure 2B**).

Young MCAO+Recan and Recan animals shared 34 DEGs, including *Aurkaip1, Pdpk1, Ndufa12, Cenpx, Med11, Spopl, Rbm6, Wtap, Usp16, Pafah1b2, Mapkap1, Derl2, Polr3k, Dusp5, D8Ertd738e, Tmem183a, Cenpv, Micu1, Slc25a4, S1pr4, Atp5f1e, Ppid, Taf9, Med21, Cep63, Mtmr14, Hmbs, Zfp367, Taf1d, Mier1, Cd3g, P2ry10, Ifi203*, and *P2ry10b*. Aged MCAO+Recan and Recan animals shared 17 DEGs, namely *Cd14, Vcan, Ly6d, Gpr18, Fcgr1, Cd79b, Mafb, Stap1, Il1r2, Cd55, Ms4a1, Ms4a6d, Cxcr5, Lrg1, Chil3, Tarm1,* and *Ngp.* Only one DEG, *Slc15a4*, was common between young and aged Recan animals (**Supplementary Figure 2D**).

GeneCards verification revealed that these DEGs included inflammation-associated genes such as *Il1r2, Cd244, Il1rn, Mmp8, Ccr2, Tnfaip2, Gsto1* (young MCAO), *Ifi203* (aged MCAO), *Cxcr2, Il1f9, Ifi203, Romo1, Pdcd5, Cd33, Hsbp1, Mmp8* (young MCAO+Recan), *Tarm1, Cxcr5, Cd79b*, and *Cd14* (aged MCAO+Recan) (30). Some of these genes, such as *Mmp8* (24 hours after onset) (57), *Ccr2* (3 days after onset) (81), and *Cxcr2* (6 hours after onset) (82), have been previously associated with ischemic stroke, but never as early as 4 hours after onset.

It is important to note that, despite the identification of 19 unique DEGs in MCAO+Recan, not all of them exhibited uniform expression trends across treatment groups. *Mrgpra2a, Mrgpra2b, Lrg1, Tarm1, Mmp8, Mt1, Hdc*, and *Pou2af1* showed sex-specific and *Cd14* showed age-specific expression patterns (|log_2_FC| ≥ 1.5, p ≤ 0.05) (**Supplementary Table 2**). The upregulation of *Cd14* post-recanalization was mainly driven by the aged group. *Mrgpra2a* and *Mrgpra2b* increased expression levels were primarily observed in female MCAO+Recan animals. Additionally, the differential changes in *Ly6d, Vcan,* and *Mt1* expression levels were observed in all subgroups within MCAO+Recan except for young males (**Supplementary Figure 2E**).

### 2. Enrichment analysis

#### i. Differentially expressed genes post-recanalization conserved across species

The PPI network of 19 DEGs of the MCAO+Recan group was constructed with STRING. The hub genes *Il1r2, Cd55, Mmp8, Cd14*, and *Cd69* were identified using the DMNC algorithm of the plug-in CytoHubba (**Figure 3A**). Since the number of DEGs in the MCAO group was low, the analysis for this group yielded no results.

**Figure 3.**
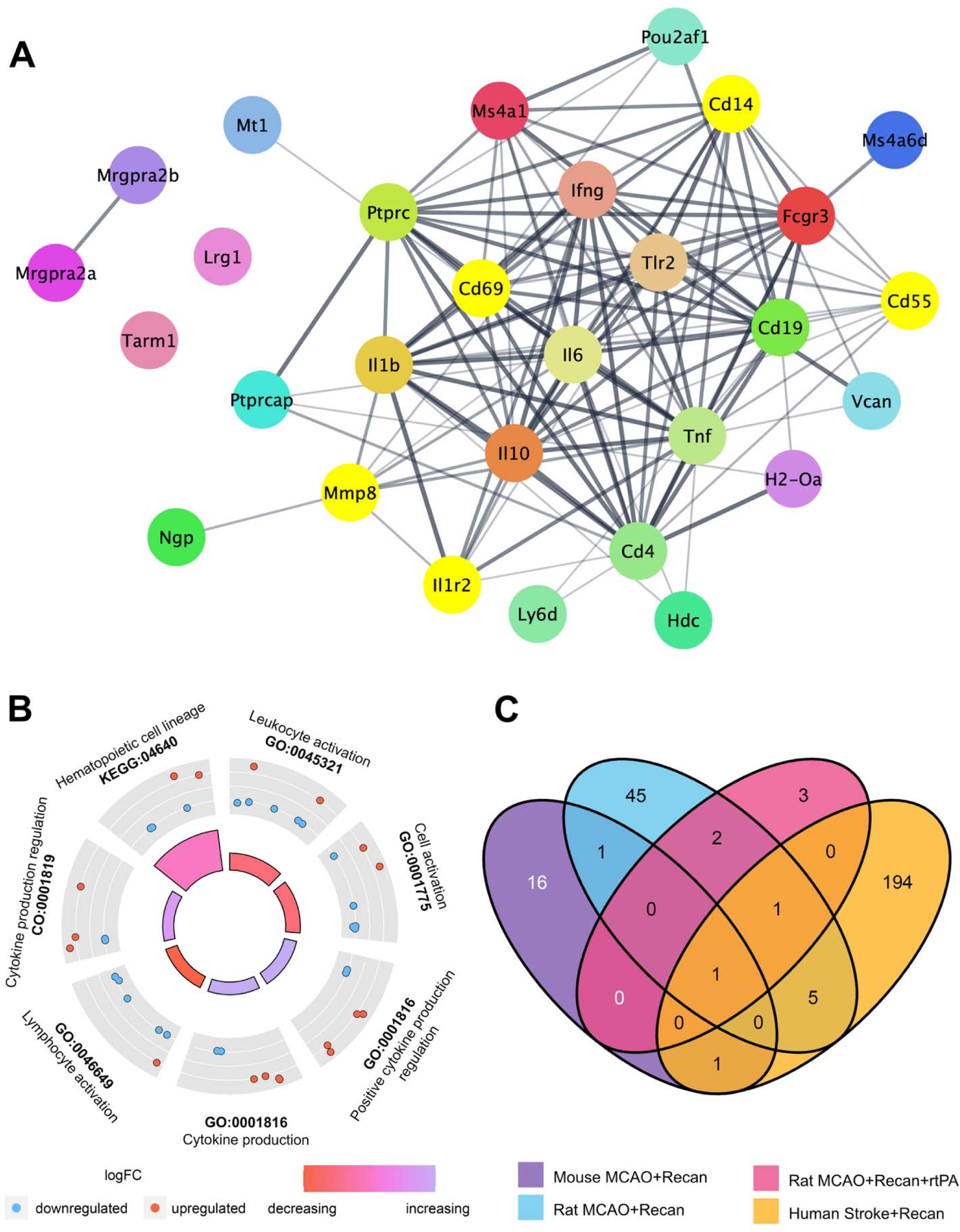
**(A)** STRING protein-protein interaction (PPI) network of differentially expressed genes (DEGs). The network was generated with a confidence level of 0.4 and a maximum of 10 additional interactors. Nodes represent DEGs, while edges indicate their interactions. Hub genes identified by the DMNC algorithm within the CytoHubba plugin are highlighted. **(B)** Circular visualization of gene annotation enrichment analysis results (GOCircle) for MCAO+Recan. Differentially expressed genes were selected (|log_2_FC| ≥ 1.5, p-value ≤ 0.05). The inner ring features a bar plot, where bar colour corresponds to the z-score (green for downregulation and yellow for upregulation), and the height of each bar represents the significance of the pathway based on log(adj p-value). The outer ring displays scatter plots of gene expression levels (logFC) for each term, with blue indicating downregulation and red indicating upregulation. **(C)** Cross-validation of all DEGs across three datasets. The analysis included mouse, GSE2113 rat samples, and GSE1656 human samples, focusing on DEGs meeting the criteria (|log_2_FC| ≥ 1.5, p-value ≤ 0.05). This comparison underscores the consistency of DEGs across different species and datasets.

To gain further biological and cell-specific context, the 19 DEGs from the MCAO+Recan group were assessed with gProfiler. The analysis showed that these DEGs were mainly involved in leukocyte activation (*Cd55, Tarm1, Mmp8, Pou2af1, Ly6d, H2-Oa, Ms4a1*), cytokine production (*Il1r2, Cd55, Tarm1, Mmp8, Pou2af1, Cd14*), and lymphocyte activation (*Cd55, Tarm1, Pou2af1, Ly6d, H2-Oa, Ms4a1*) (**Figure 3B**).

The GSE16561 and GSE21136 datasets, consisting of human and rat ischemic and recanalization samples, were used to validate the expression of the MCAO+Recan DEGs (**Figure 3C**). *Vcan* was the only DEG found in all three datasets of recanalization: GSE1656 (humans), GSE21136 (rats), and this mouse data. This suggests that *Vcan* is an important and novel marker of ischemic stroke and recanalization that is highly conserved across species. *Il1r2* and *Vcan* were shared between MCAO+Recan mice and rats. *Vcan* and *Cd14* were shared between MCAO+Recan mice and human samples. Proinflammatory *Il1r2* and *Cd14* were both hub genes, suggesting they may play a critical role in reperfusion injury and early recanalization responses.

#### ii. Sex- and age-specific inflammatory pathways post-recanalization

When breaking the MCAO+Recan enrichment analysis down to age- and sex-specific groups, many pathways are shared among the groups, and the immune system process was significantly enriched in the MCAO+Recan (**Figure 4-5; Supplementary Figure 3**). This included downregulated genes in female (*Cd55, Prg4, Ets1, Pou2af1, Cxcr5, Cd79b, Cd83, Blk, Ly6d, H2-Oa, Mzb1, Ms4a1*) (**Figure 4A**) and aged animals (*Cd55, Stap1, Cxcr5, Cd79b, Gpr18, Ly6d, Ms4a1*) (**Figure 5B**). The immune system process was also linked to upregulated genes in the female (*Cxcr2, Gpr35, Cd244a, Il36g, Il1rn, Lcn2, Il1b, Mafb, S100a8, Sirpb1a, Tlr2, S100a9, Csf3r, Bst1, Clec4d, Clec4e, Ifitm1, Nlrp12, Tarm1, Trim30b, Adam8, Ifitm2, Ifitm6, Hmox1, Hp, Mmp8, Ccr2, Alas1, Myd88, Ccr1, Tlr13, Trpm2, Lilrb4b, Lilrb4a, Osm, Wfdc17, Wfdc21, Cd300lf, Arg2, Olfm4, Acod1, Il1rap, Retnlg, App, Fpr2, Pla2g7, Trem1, Fpr1, Dusp1, Cd14, Slc15a3*) (**Figure 4A**), aged (*Gpr35, Mafb, Fcgr1, Tarm1, Ctsl, Acod1, Pram1, Cd14, Il1r2*) (**Figure 5B**), and young animals (*Cxcr2, Il36g, Il1rn, Lcn2, Hp, Mmp8, Rab27a, Pla2g7*) (**Figure 5A**). Many DEGs were shared between these groups, such as the downregulated *Cd55, Cxcr5, Cd79b, Ly6d, Ms4a1* and the upregulated *Cxcr2, Gpr35, Il1r2,* and *Arg2* (**Supplementary Table 2**).

**Figure 4.**
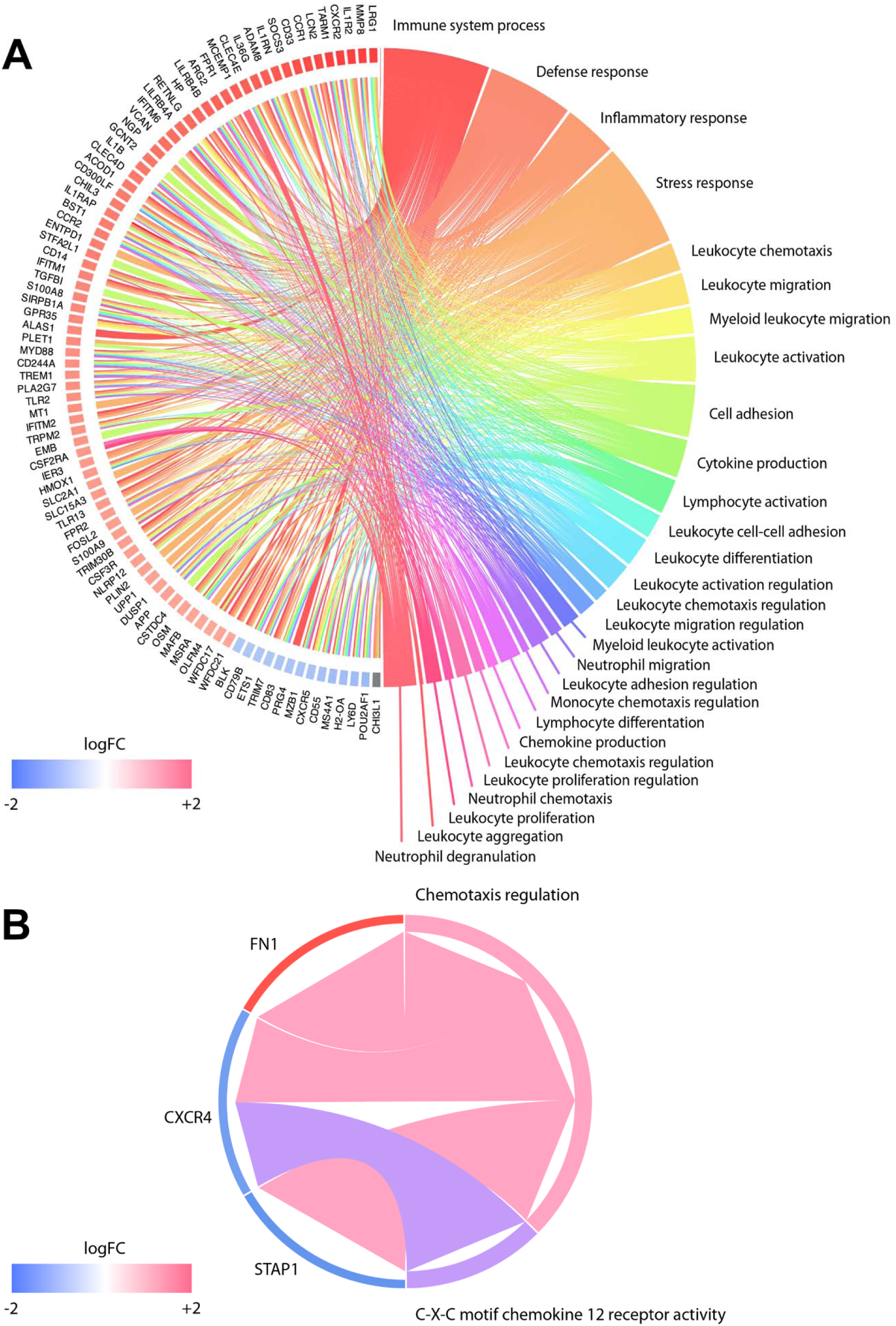
Gene ontology chord diagram illustrating core genes of significant pathways in **(A)** female and **(B)** male MCAO+Recan mice. Significant pathways are displayed on the right, while core genes are shown on the left, with their log fold change (logFC) represented by colored squares. Connections from left to right indicate gene membership within a pathway’s leading-edge subset. The gost() function was employed to analyze gene ontology using gene identifiers from the differential expression results. The analysis was conducted for Mus musculus, with parameters set to evcodes=TRUE and significant=TRUE (p-value ≤ 0.05) to highlight relevant pathways and genes.

**Figure 5.**
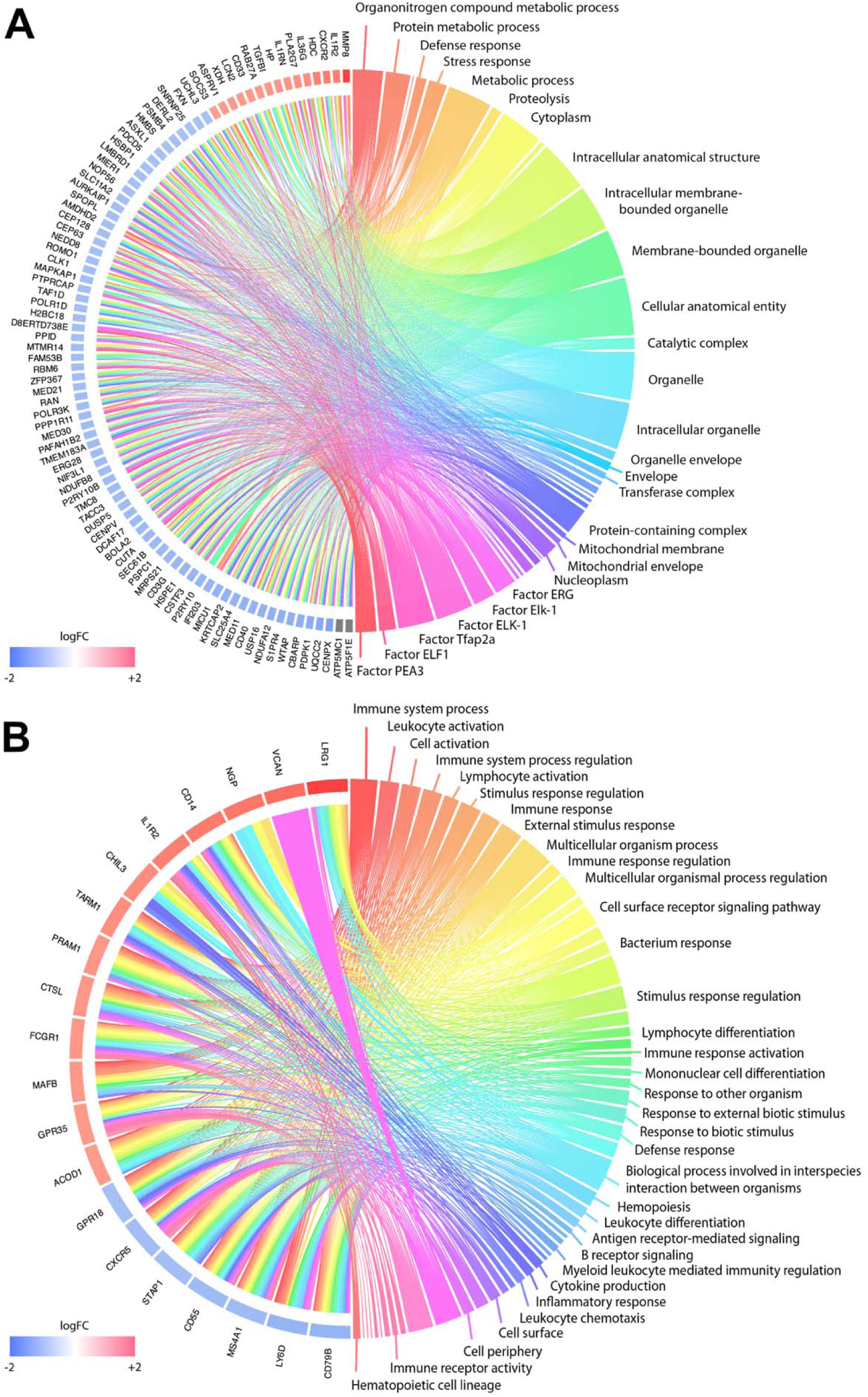
Gene ontology chord diagram showing core genes and significant pathways in **(A)** young and **(B)** aged MCAO+Recan mice. The diagram visualizes significant pathways on the right and their corresponding core genes on the left, with log fold change (logFC) indicated by coloured squares. Connections between the left (genes) and right (pathways) represent gene membership in a pathway’s leading-edge subset. The gost() function was used to analyze gene ontology, utilizing gene identifiers from differential expression results. The analysis was conducted with Mus musculus as the organism, specifying evcodes=TRUE and significant=TRUE (p-value ≤ 0.05) to highlight relevant pathways and genes.

Females and young MCAO+Recan also saw an enrichment in cytokine production and regulation. These pathways were driven by upregulated *Il1r2, Cxcr2, Gpr35, Chil1, Il1rn, Il1b, Tlr2, Csf3r, Ifitm1, Nlrp12, Ifitm2, Ifitm6, Ccr2, Myd88, Ccr1, Il13ra1, Osm, Wfdc21, Cd300lf, Socs3, Il1rap, App, Dusp1, Cd14, Csf2ra* in females (**Figure 4A**), and *Il1r2, Cxcr2, Il36g, Il1rn* in young animals (**Figure 5A**).

Other inflammatory pathways, such as lymphocyte activation and differentiation, were enriched in female and aged MCAO+Recan. Lymphocyte activation was driven by downregulated *Cd55, Cxcr5, Cd79b, Gpr18, Ly6d, Ms4a1* in the aged group (**Figure 5B**) and upregulated *Cd244a, Il1b, Mafb, Sirpb1a, Clec4d, Clec4e, Tarm1, Adam8, Ccr2, Myd88, Lilrb4b, Lilrb4a, Arg2* in the female group (**Figure 4A**). On the other hand, lymphocyte differentiation was observed in the female group with downregulated *Pou2af1, Cd79b, Cd83, Ly6d, H2-Oa, Ms4a1* (**Figure 4A**) and the aged group (**Figure 5B**) with the same genes and an additional *Gpr18*. Mononuclear cell differentiation and migration was significantly enriched in downregulated aged MCAO+Recan (*Cd79b, Gpr18, Ly6d, Ms4a1*) (**Figure 5B**) and upregulated female MCAO+Recan (*Nlrp12, Adam8, Ccr2, Ccr1, Trpm2, Retnlg, App, Fpr2, Pla2g7, Trem1, Dusp1, Pou2af1, Cd79b, Cd83, Blk, Ly6d, H2-Oa, Ms4a1*) (**Figure 4A**). Monocellular proliferation was also found in the upregulated female MCAO+Recan (*Cd244a, Il1b, Tarm1, Ccr2, Myd88, Lilrb4b, Lilrb4a, Arg2*) (**Figure 4A**).

#### iii. Neutrophil and leukocyte-based pathways upregulated in females across species

Using ImmunCellAI-mouse algorithm, our dataset showed significant changes in macrophages (M1 and M2), neutrophils, and T-cells (Tregs, T cells CD4 T gamma delta) proportion at 3 hours after reperfusion (**Supplementary Figure 4**). However, not all these cell types were observed in human orthologs by CIBERSORTx (**Figure 6**). For instance, only neutrophils had their proportion significantly increased in MCAO+Recan group in both mouse (i.e., ImmunCellAI-mouse) and human orthologs (i.e., CIBERSORTx). Notably, the proportion of neutrophils also showed both a significant main treatment effect (adj p-value = 0.0465) and a treatment × sex interaction (adj p-value = 0.0201) (**Figure 6**; **Table 1**).

**Figure 6.**
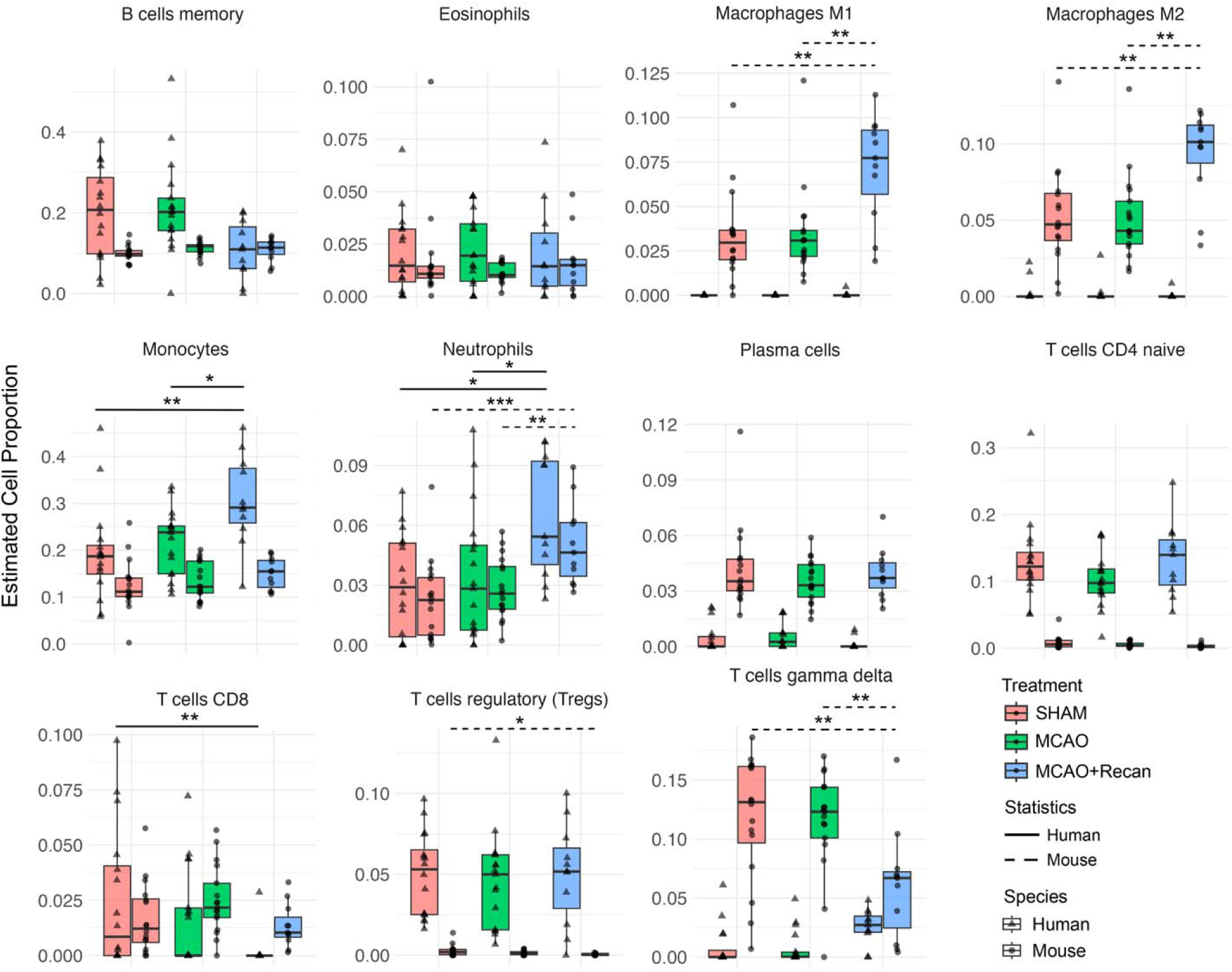
Relative immune cell proportions of cell types shared between immuCellAI-mouse (mouse reference database) and CIBERSORTx (human reference database) stratified by treatment. Results were obtained from immuCellAI-mouse and CIBERSORTx generated from microarray matrix. Statistical comparisons were performed using the Kruskal-Wallis, with p-values adjusted using the Benjamini-Hochberg method for multiple comparisons. Dunnett’s post-hoc pairwise multiple comparison was also performed (*p≤0.05, **p≤0.01, ***p≤0.001).

**Table 1.** Kruskal-Wallis and ART ANOVA analyses with Benjamini-Hochberg adjustment of CIBERSORTx and ImmunCellAI-mouse results (*adj-p≤0.05, **adj-p≤0.01, ***adj-p≤0.001).

| Cell Type | Species | Treatment effect<br>Kruskal-Wallis<br>p-val [adj p] | Treatment:Species<br>ART ANOVA<br>p-val [adj p] | Treatment:Sex<br>ART ANOVA<br>p-val [adj p] | Treatment:Age<br>ART ANOVA<br>p-val [adj p] | Treatment:Age:Sex<br>ART ANOVA<br>p-val [adj p] |
| --- | --- | --- | --- | --- | --- | --- |
| B cells naive | Human | 0.0448 [0.1112] |  | 0.6807 [0.9621] | 0.7923 [0.9640] | 0.1655 [0.4966] |
| B cells memory | Human | 0.0249 [0.0801] | **\*0.0053**<br>**[0.0175]** | 0.5132 [0.8436] | 0.0786 [0.3255] | 0.5955 [0.9010] |
| B cells memory | Mouse | 0.1187 [0.2220] |  | 0.9097 [0.9887] | 0.5952 [0.9010] | 0.1919 [0.5346] |
| Plasma cells | Human | 0.2910 [0.4220] | 0.1620<br>[0.2700] | 0.8301 [0.9857] | 0.8508 [0.9857] | 0.5783 [0.8984] |
| Plasma cells | Mouse | 0.7324 [0.8330] |  | 0.4892 [0.8395] | 0.0400 [0.2048] | 0.9145 [0.9887] |
| T cells CD8 | Human | 0.0309 [0.0899] | 0.2639<br>[0.3299] | 0.5374 [0.8579] | 0.6357 [0.9454] | 0.9454 [0.9621] |
| T cells CD8 | Mouse | 0.1274 [0.2310] |  | 0.8430 [0.9857] | 0.7537 [0.9621] | 0.9622 [0.9887] |
| T cells CD4 naive | Human | 0.2541 [0.3780] | **\*0.0089**<br>**[0.0223]** | 0.1526 [0.4830] | 0.9830 [0.9887] | 0.7488 [0.9621] |
| T cells CD4 naive | Mouse | 0.1994 [0.3212] |  | **\*\*0.0002 [0.0051]** | 0.2252 [0.5724] | 0.1736 [0.5119] |
| T cells CD4 memory resting | Human | 0.7936 [0.8389] |  | 0.3720 [0.7526] | 0.5739 [0.8984] | 0.9145 [0.9887] |
| T cells CD4 memory activated | Human | 0.0416 [0.1096] |  | 0.3238 [0.6787] | 0.1856 [0.5295] | 0.3018 [0.6483] |
| T cells follicular helper | Human | 0.4169 [0.5256] |  | **\*\*\*7.2266e-08**<br>**[3.1436e-06]** | **\*\*\*1.1697e-06**<br>**[3.3922e-05]** | **\*\*\*1.3801e-07**<br>**[4.8028e-06]** |
| T cells regulatory (Tregs) | Human | 0.7955 [0.8389] | 0.2234<br>[0.3192] | 0.9781 [0.9887] | 0.9750 [0.9887] | 0.9677 [0.9887] |
| T cells regulatory (Tregs) | Mouse | 0.0654 [0.1375] |  | 0.0346 [0.1882] | 0.0060 [0.0563] | 0.1121 [0.4178] |
| T cells gamma delta | Human | **\*0.0105 [0.0485]** | **\*\*\*1.3067e-06**<br>**[4.7912e-06]** | 0.2360 [0.5785] | 0.2544 [0.5831] | 0.0264 [0.1532] |
| T cells gamma delta | Mouse | **\*0.0109 [0.0485]** |  | 0.9590 [0.9887] | 0.6615 [0.9591] | 0.1436 [0.4812] |
| NK cells resting | Human | 0.0195 [0.0706] |  | 0.7378 [0.9621] | 0.9829 [0.9887] | 0.4261 [0.7857] |
| NK cells activated | Human | **\*0.0033 [0.0465]** |  | 0.4137 [0.7857] | 0.0103 [0.0852] | 0.0140 [0.0955] |
| Monocytes | Human | **\*0.0077 [0.0465]** | **\*0.0236**<br>**[0.0471]** | 0.0148 [0.0955] | 0.0109 [0.0866] | 0.7202 [0.9621] |
| Monocytes | Mouse | 0.2317 [0.3536] |  | 0.8165 [0.9857] | 0.1582 [0.4830] | 0.1198 [0.4255] |
| Macrophages M0 | Human | 0.6082 [0.7055] |  | 0.2232 [0.5724] | 0.0420 [0.2088] | 0.0759 [0.3255] |
| Macrophages M1 | Human | 0.2231 [0.3498] | **\*\*\*3.3698e-08**<br>**[3.3698e-07]** | **\*\*\*4.8272e-12**<br>**[3.7417e-10]** | **\*\*\*6.4512e-12**<br>**[3.7417e-10]** | **\*\*\*2.7188e-12**<br>**[3.7417e-10]** |
| Macrophages M1 | Mouse | **\*0.0057 [0.0465]** |  | 0.2270 [0.5724] | **\*\*0.0004 [0.0086]** | **\*0.0011 [0.0174]** |
| Macrophages M2 | Human | 0.7533 [0.8389] | **\*\*\*9.7579e-07**<br>**[4.8789e-06]** | 0.0061 [0.0563] | 0.0128 [0.0938] | **\*0.0028 [0.0305]** |
| Macrophages M2 | Mouse | **\*0.0080 [0.0465]** |  | 0.0314 [0.1761] | **\*0.0016 [0.0201]** | **\*0.0012 [0.0174]** |
| Dendritic cells resting | Human | 0.5842 [0.6915] |  | 0.3957 [0.7737] | 0.2014 [0.5456] | 0.2083 [0.5491] |
| Dendritic cells activated | Human | 0.1859 [0.3080] |  | 0.5327 [0.8579] | 0.0667 [0.3138] | 0.5051 [0.8436] |
| Mast cells resting | Human | **\*0.0008 [0.0243]** |  | 0.7137 [0.9621] | 0.9129 [0.9887] | 0.0767 [0.3255] |
| Mast cells activated | Human | 0.0343 [0.0948] |  | 0.4769 [0.8357] | 0.5058 [0.8436] | 0.0369 [0.1946] |
| Eosinophils | Human | 0.8616 [0.8923] | 0.8171 [0.8171] | 0.8752 [0.9887] | 0.9154 [0.9887] | 0.5232 [0.8509] |
| Eosinophils | Mouse | 0.9028 [0.9187] |  | 0.9442 [0.9887] | 0.1438 [0.4812] | 0.3208 [0.6787] |
| Neutrophils | Human | 0.0275 [0.0841] | 0.6477 [0.7197] | 0.0129 [0.0938] | 0.3844 [0.7688] | 0.4188 [0.7857] |
| Neutrophils | Mouse | **\*0.0025 [0.0465]** |  | **\*0.0017 [0.0201]** | 0.2743 [0.6119] | 0.9912 [0.9912] |
| B cells | Mouse | **\*0.0059 [0.0465]** |  | 0.6536 [0.9557] | 0.0716 [0.3194] | **\*0.0032 [0.0329]** |
| Dendritic cells | Mouse | 0.4907 [0.6055] |  | 0.3932 [0.7737] | 0.1128 [0.4178] | 0.1936 [0.5346] |
| Granulocytes | Mouse | 0.0181 [0.0699] |  | 0.4290 [0.7857] | 0.7830 [0.9621] | 0.7280 [0.9621] |
| Macrophages | Mouse | **\*0.0068 [0.0465]** |  | 0.1098 [0.4178] | **\*\*0.0005 [0.0093]** | **\*\*0.0010 [0.0174]** |
| NK cells | Mouse | 0.3161 [0.4472] |  | 0.7293 [0.9621] | 0.4064 [0.7856] | 0.1007 [0.3981] |
| T cells | Mouse | **\*0.0068 [0.0465]** |  | 0.4921 [0.8395] | 0.5139 [0.8436] | 0.2341 [0.5785] |
| T cells CD4 | Mouse | 0.1394 [0.2378] |  | **\*0.0015 [0.0195]** | 0.5455 [0.8629] | 0.2486 [0.5831] |
| NKT | Mouse | 0.1386 [0.2378] |  | 0.0566 [0.2738] | 0.1824 [0.5289] | 0.0228 [0.1367] |
| B1 cells | Mouse | **\*0.0007 [0.0244]** |  | 0.2967 [0.6454] | 0.2038 [0.5456] | 0.2547 [0.5831] |
| Follicular B cells | Mouse | 0.0143 [0.0593] |  | 0.7046 [0.9621] | 0.8536 [0.9857] | 0.1301 [0.4527] |
| Germinal center B cells | Mouse | 0.5122 [0.6189] |  | 0.6341 [0.9454] | 0.7613 [0.9621] | 0.2924 [0.6440] |
| Marginal zone B cells | Mouse | 0.0246 [0.0801] |  | 0.6681 [0.9607] | 0.4803 [0.8357] | 0.3307 [0.6799] |
| cDC1 | Mouse | 0.3748 [0.4970] |  | 0.0932 [0.3772] | 0.2439 [0.5813] | 0.7839 [0.9621] |
| cDC2 | Mouse | 0.9410 [0.9410] |  | 0.6839 [0.9621] | 0.1581 [0.4830] | 0.0696 [0.3185] |
| MoDC | Mouse | 0.0891 [0.1723] |  | 0.6506 [0.9557] | 0.9558 [0.9887] | 0.2413 [0.5813] |
| pDC | Mouse | **\*0.0102 [0.0485]** |  | 0.1176 [0.4255] | 0.0170 [0.1056] | 0.1545 [0.4830] |
| Basophils | Mouse | 0.0647 [0.1375] |  | 0.2641 [0.5969] | 0.7626 [0.9621] | 0.7644 [0.9621] |
| Mast cells | Mouse | 0.0664 [0.1375] |  | 0.8909 [0.9887] | 0.8438 [0.9857] | 0.4675 [0.8300] |
| CD4 Tm cells | Mouse | 0.0868 [0.1723] |  | 0.0144 [0.0955] | 0.4404 [0.7937] | 0.1502 [0.4830] |
| T cells helper | Mouse | 0.3771 [0.4970] |  | 0.0093 [0.0809] | 0.7663 [0.9621] | 0.3321 [0.6799] |
| CD8 Tc | Mouse | 0.7713 [0.8389] |  | 0.7852 [0.9621] | 0.8554 [0.9857] | 0.6969 [0.9621] |
| CD8 Tcm | Mouse | 0.0472 [0.1112] |  | 0.8325 [0.9857] | 0.7635 [0.9621] | 0.9682 [0.9887] |
| CD8 Tem | Mouse | 0.0479 [0.1112] |  | 0.7152 [0.9621] | 0.5953 [0.9010] | 0.4207 [0.7857] |
| CD8 Tex | Mouse | 0.3906 [0.5034] |  | 0.4425 [0.7937] | 0.9816 [0.9887] | 0.9612 [0.9887] |
| T cells CD8 naive | Mouse | 0.3458 [0.4776] |  | 0.8699 [0.9887] | 0.9270 [0.9887] | 0.9340 [0.9887] |

ART ANOVA identified significant treatment effects that differ between human and mouse for B cells memory (adj p-value = 0.0175), T cells CD4 naïve (adj p-value = 0.0223), T cells gamma delta (adj p-value = 4.7912e-06), monocytes (adj p-value = 0.0471), and macrophages M1 (adj p-value = 3.3698e-07) and M2 (adj p-value = 4.8789e-06). In humans, the proportion of monocytes showed a significant treatment effect (adj p-value = 0.0465), with the highest proportion observed in the MCAO+Recan group. However, this trend was not seen in mice (**Figure 6**). These findings validate previous observations of differing immune cell proportions between mice and humans (83,84).

Furthermore, several cell types showed significant age- and sex-specific differences. For example, macrophages M1 in mice showed not only substantial treatment effects but also treatment × age (adj p-value = 0.0086) and treatment × age × sex interactions (adj p-value = 0.0174), which aligns with human data and further emphasizes their importance. Macrophages M2 showed a similar pattern (**Table 1**; **Supplementary Figure 4**). Follicular helper T cells were notable for significant changes across all interaction terms in the human data. B cells in mice had a substantial treatment × sex × age interaction. T cells CD4 showed a significant treatment × sex interaction (**Table 1**).

The Cell Marker 2.0 database was used to identify cell types in which the DEGs were enriched. This approach provided valuable tissue and cell-specific context for the observed transcriptional changes. In this dataset, the marker with the highest gene count was for neutrophils, with all genes being significantly upregulated and driven by the MCAO+Recan group, and the greatest expression in female and aged mice (**Figure 7A**). This suggests that neutrophil markers were highly expressed and contributed to ischemic pathogenesis post-recanalization. Downregulated DEGs were mostly markers of macrophages, lymphocytes, T cells, B cells, and arterial cells (**Figure 7A**).

**Figure 7.**
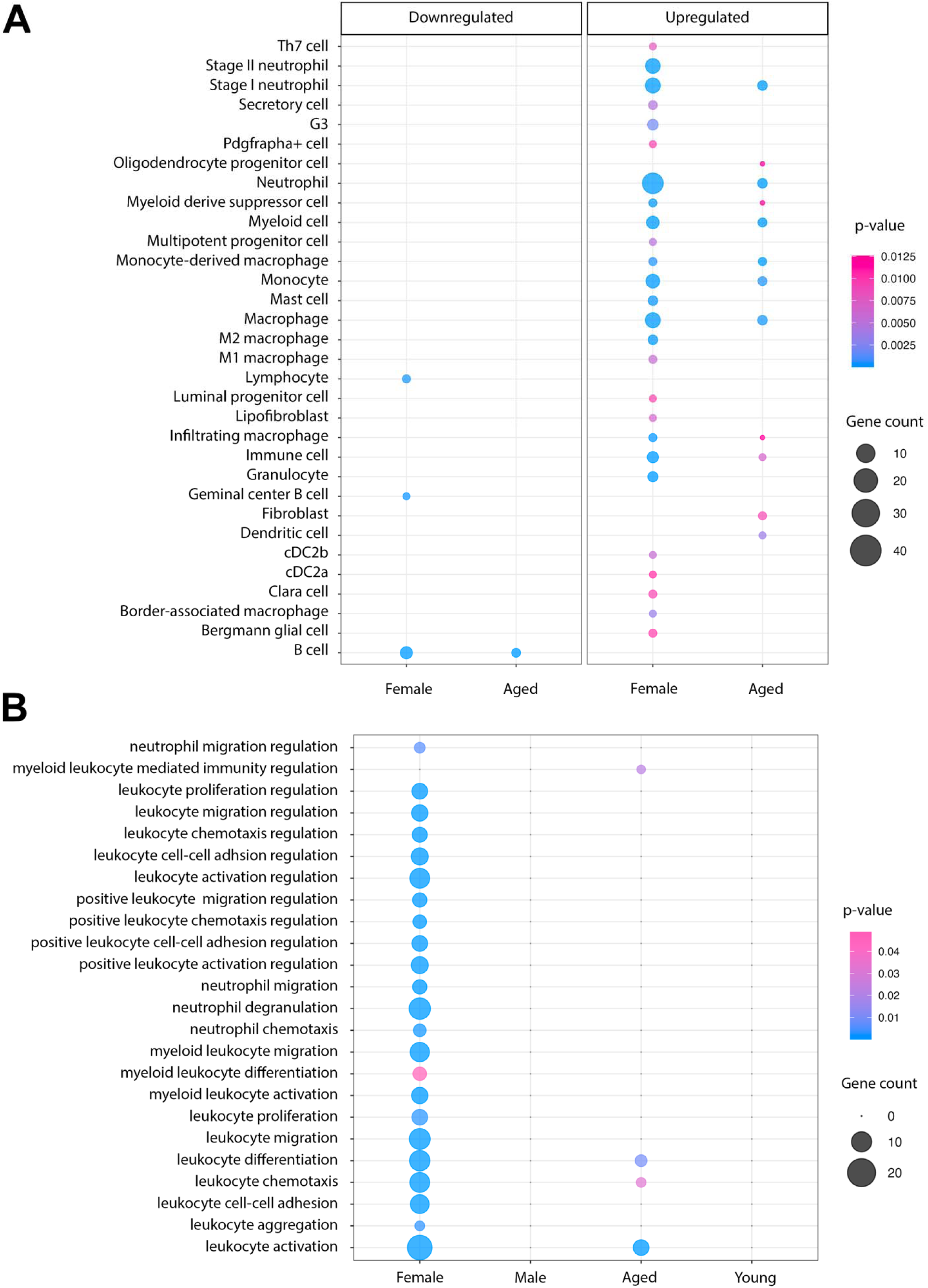
**(A)** Over-representation of cell markers from MCAO+Recan differentially expressed genes (DEGs). The Cell Marker 2.0 database and the clusterProfiler package were used to infer the cell types in which the identified DEGs might be expressed. The dot plot shows upregulated and downregulated DEGs mapped to specific cell types. **(B)** Enrichment of leukocyte and neutrophil activities in female and aged MCAO+Recan mice. Gene enrichment analysis was conducted using the gProfiler package. The enrichplot::dotplot function was used to visualize enriched pathways, with colour intensity indicating the significance of enrichment and dot size representing the gene count for MCAO+Recan samples. These highlight the significant involvement of leukocyte and neutrophil activities in the ischemic stroke model.

The female MCAO+Recan also showed the highest number of enriched pathways related to neutrophil activity, such as neutrophil migration (*Cxcr2, Il1b, Bst1, Adam8, Myd880*), chemotaxis (*Cxcr2, Il1b, S100a8, S100a9, Csf3r, Bst1, Trem1*), aggregation (*Il1b, S100a8, S100a9, Adam8*), and degranulation (*Cxcr2, Chil1, Cd55, Lcn2, S100a8, Sirpb1a, S100a9, Clec4d, Tarm1, Cd33, Adam8, Mcemp1, Hp, Mmp8, Trpm2, Lilrb4b, Lilrb4a, Olfm4, Fpr2, Fpr1, Cd14*) (**Figure 7B**).

Other potential pathways that may connect to reperfusion injury and futile recanalization in this group were leukocyte activation (*Cxcr2, Cd244a, Cd55, Il1b, Mafb, Sirpb1a, Tlr2, Clec4d, Clec4e, Tarm1, Adam8, Hmox1, Mmp8, Pou2af1, Ccr2, Cxcr5, Myd88, Lilrb4b, Lilrb4a, Cd79b, Cd300lf, Arg2, Cd83, Blk, Ly6d, App, H2-Oa, Mzb1, Ms4a1), leukocyte cell-cell adhesion (Cd244a, Cd55, Il1b, S100a8, Sirpb1a, S100a9, Tarm1, Adam8, Ets1, Ccr2, Lilrb4b, Lilrb4a, Arg2, Cd83, H2-Oa, Lrg1*), and leukocyte proliferation (*Cd244a, Cd55, Il1b, Tarm1, Ccr2, Myd88, Lilrb4b, Lilrb4a, Arg2, Blk, Mzb1, Csf2ra*). In addition, significant leukocyte differentiation (*Mafb, Cd79b, Ctsl, Gpr18, Ly6d, Ms4a1*), leukocyte chemotaxis (*Gpr35, Stap1, Cxcr5, Gpr18*), and leukocyte activation (*Cd55, Mafb, Stap1, Tarm1, Cxcr5, Cd79b, Ctsl, Gpr18, Ly6d, Pram1, Ms4a1*) were found in the aged MCAO+Recan (**Figure 7B**).

The high degree of expression in neutrophil-associated pathways in these samples was then compared with other datasets related to recanalization after stroke. The female MCAO+Recan group was compared to the human female samples GSE1656 (GSE21136 was excluded as it consists of only male samples), and 37 shared orthologues, including the hub genes *Il1r2, Mmp8, Cd14*, and *Cd55,* were identified (**Figure 8A**). Notably, *Vcan* was also significantly upregulated in both female mice and female patients. Furthermore, the two groups shared a considerable number of enriched pathways (**Figure 8B**), many of which were related to neutrophil and leukocyte activity (**Figure 8C**). This indicates a highly conserved pattern of post-recanalization leukocyte- and neutrophil activities across species that can be linked to reperfusion injury.

**Figure 8.**
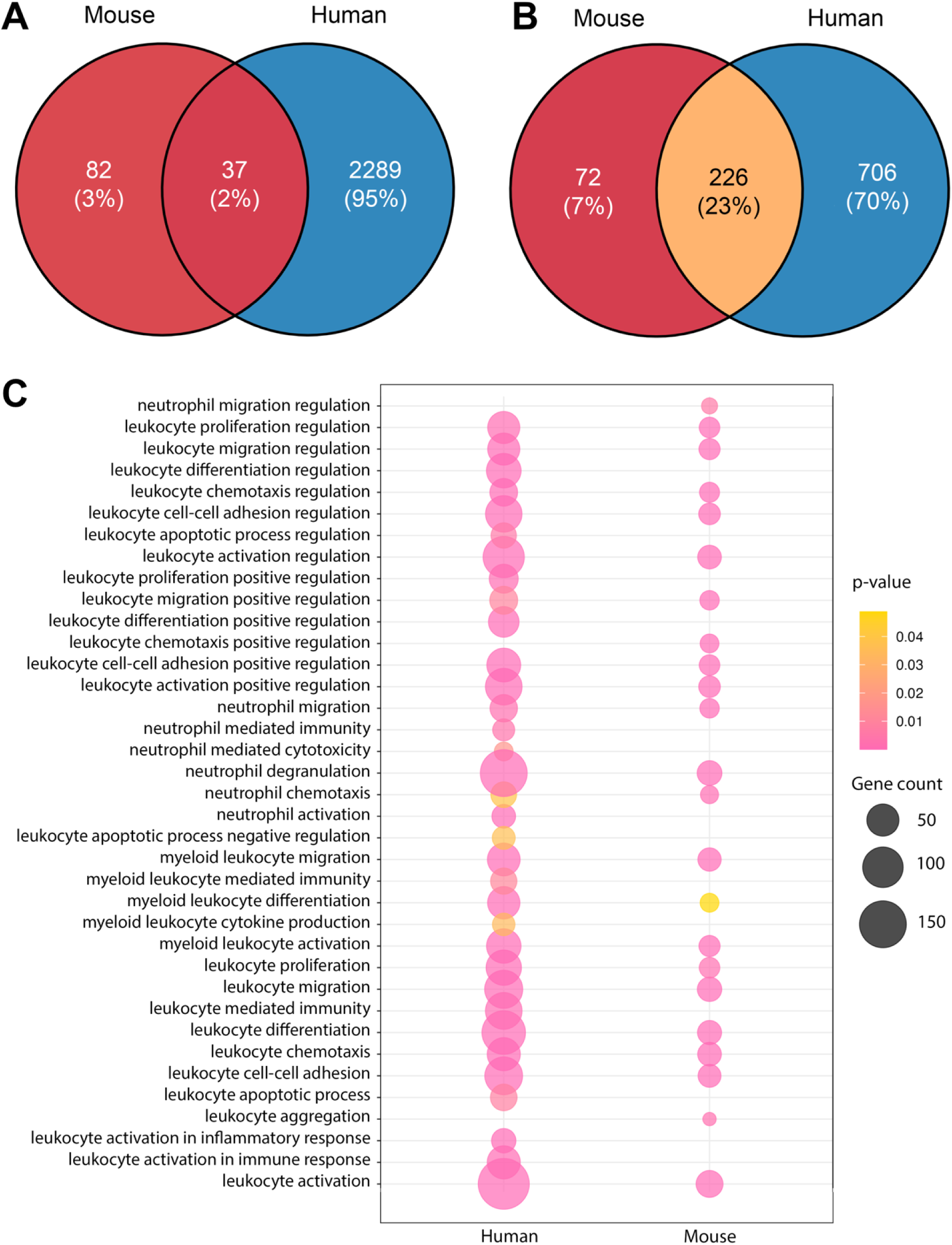
**(A)** Orthologue differentially expressed genes (DEGs) shared between MCAO+Recan female mice and clinical female patients. DEGs were identified in mouse and human datasets using the limma package (|log_2_FC| ≥ 1.5, p-value ≤ 0.05). The analysis highlights the common transcriptional responses to ischemic stroke and reperfusion (MCAO+Recan) across species. **(B)** Shared gene ontology (GO) terms and KEGG pathways between MCAO+Recan mice and clinical patients. Functional enrichment analyses were performed using the gProfiler package to identify overlapping biological processes and pathways. Key shared pathways related to stroke, inflammation, and immune responses were identified, showing conserved functional responses across the species. **(C)** Conservation of leukocyte- and neutrophil-based pathways in human and mouse samples. Pathways related to leukocyte and neutrophil activity were conserved between human clinical patients and MCAO+Recan mice.

### 3. Regulatory network analysis

Using NetworkAnalyst, the miRNAs that regulated DEGs in the MCAO+Recan group were predicted. The top regulated nodes (degree ≥ 10) were identified to be *Clec7a, Cd69, Ets1, Cxcr2, Macf1, Csf3r, Csf1, Btc, Vcan, Entpd1, Slc11a2, Cxcr2, Fitm2, Lmbrd1, Pafah1b2*, and *Gsk3b* (**Supplementary Figure 6)**.

Notably, *Vcan* was the sole node identified in aged MCAO+Recan animals and one of the central nodes in female MCAO+Recan animals. It was regulated by mmu-mir-450b-5p, mmu-mir-143-3p, mmu-mir-340-5p, mmu-mir-680, mmu-mir-690, mmu-mir-185-5p, mmu-mir-203-5p, mmu-mir-129-5p, mmu-mir-3079-5p, mmu-mir-434-3p in the aged group, and the additional mmu-mir-203-5p and mmu-mir-185-5p in female group (**Supplementary Figure 6**). *Vcan* was an upregulated gene involved in the cell adhesion pathway and cell periphery in females and in the extracellular region for both the female and aged groups. It is also regulated by the NF-κB factor (motif: GGGRATTTCC) (**Supplementary Table 2**). This suggests that *Vcan* plays a role in remodelling the extracellular matrix (ECM) during neutrophil migration and adhesion in reperfusion injury.

The top 50 TFs that regulated DEGs in MCAO+Recan groups were exported from ChEA3. NFE4 and MTF1 had the highest confidence score, 9.0 and 10.0, respectively, and regulated genes in females from the MCAO+Recan group **(Supplementary Figure 7)**. NFE4 regulated mostly proinflammatory genes, namely *P2ry13, Ifitm2, Csf3R, Fpr1, Mcemp1, Fpr2, Csf2Ra, Trem1, Bst1, Nlrp12, Clec4D, Cxcr2, Adam8, Clec4E, Cd300Lf, Myd88, Cd33, Ccr2*, and *Tlr2*. MTF1 targeted more specific leukocyte differentiation and leukocyte aggregation and migration, including *Il1rn, Csf3R, Fpr1, Tnfaip2, Fpr2, Csf2ra, Trem1, Pla2g7, Socs3, Clec7A, Cxcr2, Upp1, Cd14, Cd300lf, Slc15A3, Slc16A3, Ier3, Ccr1, Osm, Acod1, Il36G, Cyb561A3, Fosl2, Bst1, Nlrp12, Clec4D, Mafb, Il1B, Adam8, Clec4E, S100A9, Myd88*, and *Tlr2*. These two TFs co-regulated multiple genes, namely *Cd300lf, Csf2ra, Nlrp12, Clec4d, Csf3r, Trem1, Cxcr2, Adam8, Fpr1, Fpr2,* and *Bst1*.

Among the regulated genes, 16/19 of NFE4 targeted genes (*Csf3R, Fpr1, Mcemp1, Fpr2, Csf2Ra, Trem1, Bst1, Nlrp12, Clec4d, Cxcr2, Adam8, Clec4e, Cd300lf, Myd88, Cd33, Ccr2, Tlr2*) and 17/33 of MTF1 targeted genes (*Csf3R, Fpr1, Tnfaip2, Fpr2, Csf2Ra, Trem1, Pla2G7, Socs3, Clec7A, Cxcr2, Upp1, Cd14, Cd300lf, Slc15A3, Slc16A3, Ier3, Ccr1, Osm, Acod1, Il36g, Cyb561A3, Fosl2, Bst1, Nlrp12, Clec4d, Mafb, Il1b, Adam8, Clec4e, S100A9, Myd88, Tlr2*) were implicated in neutrophil migration, degranulation, chemotaxis, and migration, both involved in neutrophil migration and degranulation (**Figure 4A; Supplementary Table 2**). Combining these multifaceted data, a network illustrating how DEGs, miRNAs, and TFs interact to upregulate neutrophil activity, implicating reperfusion injury, particularly in females, was constructed (**Figure 9**).

**Figure 9.**
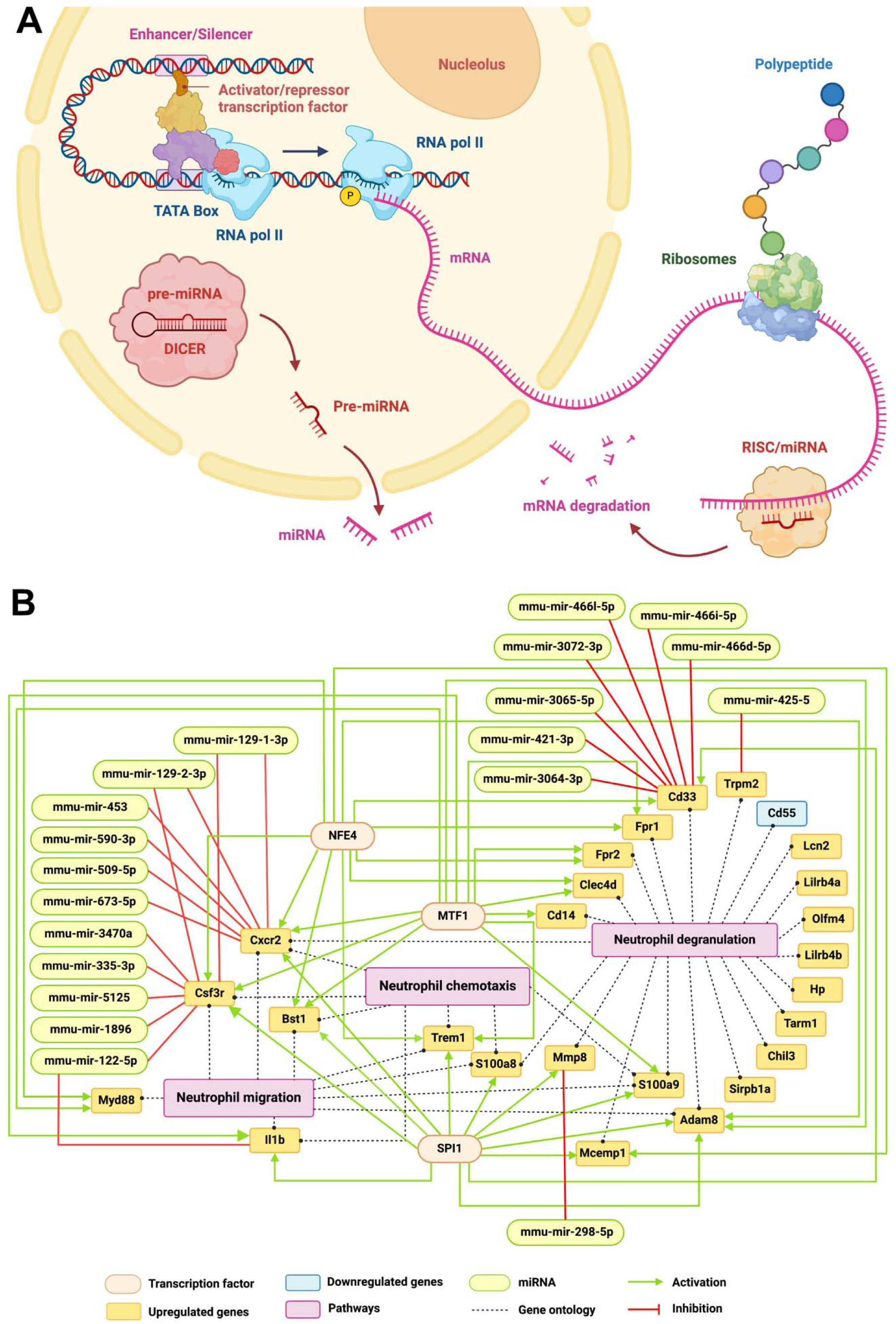
**(A)** Transcriptional regulation by transcription factors (TFs) and miRNAs. mRNA is transcribed from DNA in the nucleus by the DNA polymerase II complex, a process regulated by TFs, which can act as activators or repressors. TFs bind to specific DNA regions, either enhancers or silencers, to increase or decrease gene transcription. In the cytoplasm, pre-miRNAs mature into miRNAs, which can cleave mRNA strands, destabilize mRNA, or inhibit translation. **(B)** Neutrophil signalling network. The neutrophil signalling network was generated using integrated results from gProfiler enrichment, NetworkAnalyst, and ChEA3 analyses. One TF can regulate several genes, and multiple TFs may regulate a single gene. Similarly, multiple miRNAs can regulate one gene, and each miRNA can target more than one gene. Graphs were created with BioRender.

## Discussion

### 1. Novel biomarkers of recanalization and reperfusion failure

Several blood biomarkers have been identified with high sensitivity for stroke diagnosis, such as S100B (glial activation), vWF (thrombosis), MMP9, and VCAM (inflammation), and are used to discriminate stroke from healthy controls (85). As the number of candidate biomarkers continues to rise, they offer additional insights into clinical responses to medical interventions and outcome predictions. However, there are currently no biomarkers for the early prediction of the risks of poor recovery following recanalization in ischemic stroke. The requirements for such biomarkers, including high sensitivity, specificity, and rapid turnaround time, make their development challenging. Identifying blood biomarkers could revolutionize stroke care, enhancing prognosis and recovery with personalized treatment. At-risk patients can then benefit from early additional therapies such as neuroprotective or anti-inflammatory. Moreover, early biomarkers could help stratify patients in clinical trials, offering better insights into the mechanisms of injury and recovery. The quest for reliable biomarkers is, therefore, critical to improving post-recanalization outcomes and reducing long-term disability in stroke survivors.

This study investigated the regulation of genes associated with ischemia and recanalization by transcription factors and miRNAs, uncovering early changes associated with neutrophil activity that may drive pathophysiology related to reperfusion injury and futile recanalization. In the co-regulatory network, several nodes exhibited high centrality, including five hub genes—*Il1r2, Cd55, Mmp8, Cd14, and Cd69*—along with two transcription factors, MTF1 and NFE4, and multiple miRNAs. These regulatory elements were predominantly associated with neutrophil activity, offering a deeper understanding of gene regulatory mechanisms involved in reperfusion injury and recovery post-stroke and recanalization.

Further analysis of the hub genes suggests *Il1r2* expression may play a regulatory role in modulating the inflammatory response by reducing IL-1 activity, potentially influencing stroke outcomes by limiting excessive inflammation (86). *Cd55*, downregulated following recanalization, can protect brain tissue in ischemic stroke by inhibiting the complement system, preventing excessive inflammation and complement-mediated damage to neurons, glial cells, and the blood-brain barrier. This regulation helps preserve tissue integrity and reduce secondary injury following the initial ischemic event (87–89). *Mmp8*, primarily produced by neutrophils, facilitates neutrophil migration and plays a role in degrading the ECM components, thereby modulating inflammation. *Mmp8* expression is associated with high risks of ischemia, contributes to the breakdown of the blood-brain barrier and tissue remodelling, and decreased MMP8 protein levels improve neurological functions and reduce MCAO infarct size in rats (57,90). Additionally, *Cd69* and *Cd14* are integral components of the core inflammatory program governing neutrophil activity during ischemic stroke. For instance, *Cd69* dampens endothelial activation during ischemic events, and its deficiency in mouse models is linked to increased neutrophil infiltration and larger infarct volumes (91). *Cd14*, on the other hand, triggers neutrophil responses through toll-like receptors, amplifying the inflammatory response. Elevated *Cd14* expression has been associated with worse stroke outcomes due to increased inflammation (92). Notably, a limitation of this study is that its design cannot distinguish between transcriptional changes driven by recanalization and/or reperfusion injury or whether they primarily reflect the length of time (i.e., 4 hours) from ischemic onset.

miRNAs, which directly influence mRNA expression, also play a crucial role in this regulatory network. The most connected genes in this miRNA analyses were *Clec7a, Cd69, Ets1, Cxcr2, Macf1, Csf3r, Csf1, Btc, Vcan, Entpd1, Slc11a2, Cxcr2, Fitm2, Lmbrd1, Pafah1b2*, and *Gsk3b*. The genes *Cxcr2, Vcan, Gsk3b, Csf3r*, and *Ets1* were particularly relevant to neutrophil migration and ECM remodelling, the critical processes of ischemic stroke. For instance, CXCR2 is a chemokine receptor critical for neutrophil migration. It binds to chemokines like CXCL1 and CXCL8, directing neutrophils to sites of inflammation during ischemic stroke. CXCR2 has a direct role in recruiting neutrophils to damaged brain tissue, contributing to inflammation and stroke pathology (13). CSF3R is the receptor for granulocyte colony-stimulating factor, which regulates neutrophil production and release (93). The roles of other genes are less clear. GSK3B is a kinase involved in cell migration, proliferation, and apoptosis and also regulates ECM components and may therefore be involved in stroke recovery by modulating tissue remodelling (94). MACF1 is involved in cytoskeleton dynamics, essential for cell migration and structural integrity (95). Though its direct role in ischemic stroke is unclear, MACF1 could affect cell migration, including neutrophil motility and tissue repair processes.

### 2. Growing evidence of neutrophils’ central role in reperfusion injury

The functional enrichment analysis and cell deconvolution analysis here highlighted that the biological processes regulated by these genes were primarily related to leukocyte and neutrophil activity. Neutrophil microcirculatory stalls represent an emerging area of research in ischemic stroke and futile recanalization. While some *in vivo* imaging studies have shown neutrophil stalls in the microvasculature are the driving cause of microcirculatory failure, much remains to be learned (13).

The baseline immune composition differs profoundly between humans and mice, most notably in the proportions of neutrophils and lymphocytes. Humans have a neutrophil-dominant profile (50–70% of white blood cells), whereas mice are lymphocyte-rich (70–90%) with far fewer circulating neutrophils (10–30%) (96–99). These inherent disparities extend to other immune subsets, including transitional B cells, memory T cells, and macrophage populations, and must be considered when interpreting cross-species immune responses. Our transcriptomic deconvolution analysis revealed apparent species-specific treatment effects. Monocytes exhibited a significant treatment response in humans but not in mice, whereas neutrophils responded in both species, with comparable treatment-driven patterns, despite their markedly different baseline abundances. This highlights the strong potential of neutrophils for translational research in studying reperfusion injury or futile recanalization. This also suggests that the observed neutrophil-driven biomarkers have a high probability of translating to clinical studies. Several mouse-restricted subsets, such as B1 cells, pDCs, and specific macrophage subtypes, also exhibited significant treatment effects, underscoring the need for cell-type– and species–specific analyses.

After stroke, neutrophils heavily rely on the ECM to migrate, extravasate, and aggregate in small capillaries. Ischemic injury triggers inflammatory cytokines and chemokines, upregulating ICAM-1 and VCAM-1 on endothelial cells. Neutrophils bind to these adhesion molecules via integrins (e.g., α2β1, α4β1), and secrete MMP-9, degrading ECM components like collagen and fibronectin, facilitating extravasation. Once inside the brain parenchyma, neutrophils follow chemokine gradients (e.g., CXCL1) to migrate, but as they accumulate, they release neutrophil extracellular traps that trap more neutrophils and platelets, leading to microvascular plugging and obstruction, particularly in small vessels within the ischemic core and penumbra. The interaction between neutrophils and the altered ECM thus plays a crucial role in their ability to migrate and aggregate after a stroke, causing reperfusion injury and potential futile recanalization.

### 3. *Vcan* as a conserved recanalization marker between different animal models

*Vcan*, encoding the large extracellular matrix chondroitin sulfate proteoglycan versican, emerged from this study as a novel player in ECM-driven neutrophil-mediated microcirculatory failure or reperfusion injury. Interestingly, *Vcan* has not been previously investigated in the context of ischemic stroke or futile recanalization, despite this study’s observation of its significantly conserved increased expression across species that underwent recanalization treatment. In traumatic brain injury, it was shown that FOXO1 protein is induced in neutrophils after injury, and FOXO1 upregulates cytoplasmic *Vcan*. These FOXO1-high neutrophils infiltrate the brain, promoting the aggregation and activation of other immune cells, exacerbating brain damage (100). In other pathologies, *Vcan* and other ECM proteoglycans play a crucial role in ECM remodelling during recovery, interacting with proteins and molecules to maintain ECM structure and stability. *Vcan* not only acts as a biomarker of peripheral ECM remodelling but also serves as a repository for cytokines that regulate immune cell behaviour (101). These dynamic interactions between ECM and neutrophils, where neutrophils contribute to ECM remodelling via enzymes like neutrophil elastase and matrix metalloproteinases, while the ECM influences neutrophil function, highlight the bidirectional relationship that drives inflammation and recovery. *Vcan*’s role in this process emphasizes its significance in shaping the inflammatory microenvironment, influencing both neutrophil activity and ECM integrity, which are vital to stroke progression and post-reperfusion outcomes. Beyond these connections, however, little is known regarding *Vcan*’s role in stroke or recanalization outcomes, marking it as an intriguing target for further research.

Previous single-cell RNA sequencing (scRNA-seq) data identified monocytes, but not neutrophils, as the primary leukocyte population expressing *Vcan* (102). This study, however, employed a 35-minute MCAO mouse model and collected blood samples for scRNA-seq analysis at days 2 and 14 post-stroke, which differ from the model and sampling strategy used in this study. Differences in both experimental design (i.e., 35-minute MCAO being a less severe model) and temporal resolution likely contribute to the observed discrepancies. Neutrophil and monocyte dynamics after ischemic stroke follow distinct temporal patterns: neutrophil counts increase rapidly within hours, reach a peak within the first 24 hours, and decline within approximately one week, whereas monocyte counts increase more gradually and remain elevated for an extended period (103–105). These kinetics may explain why *Vcan* expression was previously associated with monocytes at later stages, while this study linked it to neutrophils at earlier time points. Together, these findings suggest that the cellular sources of *Vcan* expression may vary across different phases of ischemic stroke and reperfusion. Additional scRNA-seq studies with refined temporal sampling are required to determine whether *Vcan* expression is differentially regulated by neutrophils in the acute phase and by monocytes at later stages.

In aged animals, *Vcan* was significantly regulated by mmu-mir-450b-5p, mmu-mir-143-3p, mmu-mir-340-5p, mmu-mir-680, mmu-mir-690, mmu-mir-185-5p, mmu-mir-203-5p, mmu-mir-129-5p, mmu-mir-3079-5p, mmu-mir-434-3p, and the additional mmu-mir-203-5p and mmu-mir-185-5p for female MCAO+Recan. Some of these miRNAs already have established roles in ischemic stroke. miR-143-3p and miR-185-5p are highly sensitive markers of ischemic stroke in clinical patients (106,107). miR-143-3p plays a role in vascular remodelling, integrity, and angiogenesis by regulating vascular smooth muscle cells, which are crucial for stroke recovery and reperfusion (106). Meanwhile, miR-185-5p demonstrates neuroprotective effects in ischemic stroke by targeting genes involved in oxidative stress and apoptosis. Overexpression of miR-185-5p has been shown to protect neurons from ischemic injury by inhibiting pro-apoptotic genes like Caspase-3 and reducing oxidative stress through the NF-κB pathway (107).

Additionally, miR-450b-5p is highly expressed in thrombi retrieved via EVT from stroke patients (108), while miR-340-5p exhibits anti-inflammatory properties, inhibiting pro-inflammatory cytokines such as TNF-α and IL-1β. Overexpression of miR-340-5p in stroke models reduces infarct size and improves functional recovery by mitigating inflammation (109). miR-129-5p further enhances ischemic brain recovery by binding to SIAH1 and activating the mTOR signalling pathway, preventing neuronal apoptosis (110). Together, these findings suggest that these miRNAs, specifically those found to be modulating *Vcan*, represent potential therapeutic targets, offering promising avenues for future interventions to improve clinical outcomes following recanalization.

### 4. Key transcriptional regulators in reperfusion injury

ChEA analysis showed that NFE4 and MTF1 were the TFs with the highest scores in the entire dataset, regulating 19 and 33 genes, respectively, in the female MCAO+Recan group. Among these, 16/19 and 17/33 were implicated in neutrophil migration, degranulation, chemotaxis, and migration. The high number of genes linked to neutrophil function suggests that NFE4 and MTF1 were central regulators in mediating the recruitment and activity of neutrophils, which could profoundly affect stroke recovery and outcomes, positioning them as key players in the reperfusion injury after ischemic stroke. Previous studies demonstrated that in brain ischemia induced by transient middle cerebral occlusion, MTF-1 was triggered by a subsequent temporary femoral artery occlusion (i.e. treatment via remote ischemic conditioning), mediating neuroprotective mechanisms against oxidative and hypoxic stress. MTF-1 also translocate to the nucleus and induces NCX1 upregulation, which has been demonstrated to be neuroprotective (111). NFE4, however, has not been linked to stroke previously.

TFs function as regulators that modulate transcriptional output, thereby controlling cell fate decisions and inflammatory responses. Previous studies showed that TFs instruct a transcriptional program that allows the developmental transition from neutrophil precursors into mature neutrophils (112). However, the molecular mechanisms underlying neutrophil differentiation and function during inflammation remain largely uncharacterized. By regulating genes involved in neutrophil migration, degranulation, chemotaxis, and cellular migration, NFE4 and MTF1 likely influence the timing, intensity, and localization of neutrophil activity in the ischemic brain. For instance, genes under their control could dictate how quickly neutrophils are recruited to the injury site, the extent to which they degranulate to release enzymes that break down damaged tissue, and how they move through the damaged brain microenvironment. Such regulation is critical because an excessive or prolonged neutrophil response can lead to blood-brain barrier disruption, increased infarct size, and further neuronal death.

In contrast, a finely tuned response can promote tissue repair and resolution of inflammation. Thus, the role of NFE4 and MTF1 in modulating neutrophil-related genes highlights their importance in balancing these opposing outcomes. On one hand, they facilitate necessary immune functions that clear damaged cells and initiate healing, while on the other, they must restrain excessive inflammation to prevent further injury. The ability of these transcription factors to tightly regulate neutrophil behaviour may be critical in determining whether stroke recovery proceeds in a neuroprotective or detrimental direction, influencing both short-term recovery and long-term neurological outcomes. Understanding the precise mechanisms by which NFE4 and MTF1 control these processes could open avenues for developing targeted therapies that modulate neutrophil responses, potentially minimizing damage and enhancing recovery in stroke patients.

While ChEA effectively infers the activity of these TFs based on the enrichment of their target genes, it is important to note that this analysis does not directly indicate whether the transcription factors themselves are deregulated at the mRNA or protein expression level. Post-translational modifications, such as phosphorylation, acetylation, ubiquitination, or sumoylation can also modulate TF localization, stability, DNA-binding affinity, and interaction with co-factors, thereby fine-tuning TF regulatory functions without necessitating changes in gene expression. An altered activity inferred by ChEA could arise from changes in the candidate TF’s own expression, but it could also stem from more complex regulatory mechanisms. For instance, MTF1 is known to be activated by zinc binding and can be regulated by phosphorylation, illustrating how such modifications can dictate its functional state (113,114). Therefore, even if NFER4, MTF1, or SPI1 potentially had no significant changes in their expression, their inferred hub status through ChEA strongly suggests that their activity is altered, potentially through critical post-translational modification events. To fully ascertain their direct deregulation, further investigation into their respective mRNA expression (e.g., via RNA-seq or qRT-PCR data) and protein abundance (e.g., through Western blotting or mass spectrometry) in our specific experimental conditions would be necessary.

### Limitations

Inherent limitations, including a moderate sample size, should be considered when interpreting the findings of this study. While larger sample sizes can enhance statistical power, our study was designed as an exploratory investigation to capture early and rapid transcriptional changes following ischemia and recanalization at time points that had not previously been studied. Supporting evidence from previous studies suggests meaningful results can be obtained even with relatively small sample sizes (e.g., between 8 and 16) for microarray studies (115–117). A permutation method that accounts for correlation and effect size heterogeneity among genes shows that a small pilot dataset with 4-6 samples per group is sufficient (118). Furthermore, analyses of the main effects comparing SHAM (N=16) versus MCAO+Recan (N=11) groups were sufficiently powered to define changes in gene expression associated with stroke and recanalization. Nevertheless, more extensive studies on subgroup analyses (such as sex and age) could benefit from a larger sample size.

These findings identify early genomic indicators of ischemia and recanalization for further investigation in future studies. The data reveal consistent molecular patterns across different biological conditions that can inform age and sex-specific gene expression and be further investigated as modulators of pathology and outcome. With multiple age and sex groups, the consistency of findings across groups suggests that the observed gene expression changes are robust and meaningful, providing a strong foundation for further investigation. Given the extensive number of gene expression changes observed, comprehensively validating all differentially expressed genes within a single study would be impractical. However, our exploratory approach allowed for efficient identification of the most relevant genes for further validation using functional and outcome-related measures. Validation studies incorporating a larger sample size, additional assays of leukocyte activation and function during and after ischemia, and additional time points will further inform the functional importance of gene expression patterns and enhance translational relevance.

Our study is unable to conclusively demonstrate whether observed transcriptomic differences arise from changes in gene expression within a given cell type or from shifts in immune cell composition—such as stroke-induced leukocytosis—captured in bulk RNA-seq data. For example, increased detection of canonical neutrophil genes (e.g., *Il1r2*, *Mmp8*) shortly after ischemia may reflect higher circulating neutrophil counts rather than transcriptional activation within individual neutrophils (119–121). While our use of computational deconvolution provides an improved estimate of cell type abundance, it cannot fully resolve cell type–specific transcriptional programs. Future studies incorporating scRNA-seq or single-nucleus approaches would enable direct discrimination between compositional shifts and actual within-cell transcriptional changes, providing a more precise understanding of the molecular mechanisms driving immune responses after stroke.

Another key consideration in the study design was the selection of early time points, specifically 1 hour after ischemic onset and 3 hours after recanalization (4 hours after onset). This decision was guided by the need to capture rapid transcriptional changes within the therapeutic window, which holds the most significant potential to inform acute therapeutic strategies. Currently, intravenous thrombolysis with r-tPA is most effective within 3-4.5 hours of stroke onset, while mechanical thrombectomy benefits select patients for up to 24 hours based on advanced imaging criteria (18,19,122,123). Later time points, such as 24 hours post-recanalization, were not included because they fall outside the window for meaningful acute interventions and may primarily reflect secondary injury processes rather than immediate molecular responses. Identifying the earliest transcriptional responses is, therefore, crucial for developing interventions that can be administered adjunctively with recanalization therapies to prevent irreversible neuronal loss and enhance functional recovery. Moreover, previous research has demonstrated that gene expression changes can occur as early as 15 minutes after stimulation, reinforcing the relevance of capturing early-phase transcriptional dynamics (25). Neutrophils are known to respond rapidly to ischemic and inflammatory cues, making this timeframe appropriate for examining their involvement in stroke microcirculatory failure and futile recanalization (13,24). Future studies could incorporate time-course analyses extending beyond the acute phase to determine whether early gene expression changes translate into sustained neuroprotection or contribute to delayed injury mechanisms.

The study did not compare gene expression between venous and arterial blood samples but rather focused on peripheral transcriptional changes in venous blood. Arterial blood, being directly oxygenated and closer to the site of cerebral events, may exhibit different gene expression patterns than venous blood (124,125). Moreover, arterial blood reflects systemic physiological responses and oxygenation status more directly, while venous blood may better capture localized tissue responses and metabolic byproducts (126). Studying peripheral changes is especially valuable in clinical settings where rapid, minimally invasive, and repeatable sampling is required. Unlike arterial sampling, which is more invasive and technically challenging, venous blood draws are routinely performed and can facilitate the identification of early biomarkers predictive of stroke outcomes. Nevertheless, future research could explore potential differences in transcriptional profiles between venous and arterial blood to determine their relative utility in biomarker discovery and stroke prognosis.

The absence of behavioural or functional outcome in this study does not allow direct correlation of behaviour with gene expression changes. This study was not designed as an outcome-based investigation but rather as an exploratory study to identify early molecular patterns following recanalization. By prioritizing early transcriptional changes, our study provides a necessary first step in understanding the mechanisms that drive post-stroke repair. This allows future research to take a more targeted approach to investigate the functional impacts of individual therapeutic targets (genes and gene products), focusing on the most promising molecular pathways identified here. This study provides a strong foundation by identifying key gene expression changes that occur immediately after recanalization, offering potential early indicators of stroke recovery that can be explored in subsequent studies incorporating longitudinal behavioural measures, imaging modalities, and long-term follow-up such as with the use of knock-out/knock-in models.

## Conclusion

This study illuminates the critical role of gene regulatory networks in the pathophysiology of ischemic stroke, particularly in early recanalization responses. By identifying essential hub genes—*Il1r2, Cd55, Mmp8, Cd14,* and *Cd69*—and the novel marker *Vcan*, this study uncovered genomic indicators of recanalization that are highly conserved across species, from mice to humans. *Vcan* presents a promising target for future investigation, as its interaction with neutrophils during microcirculatory stalls and a high degree of conservation across species may offer new therapeutic avenues. Neutrophil was the only cell type with deconvoluted cell proportion significantly increased in both human and murine reference data. The functional enrichment analysis also revealed that pathways related to leukocyte migration, neutrophil activation, and phagocytosis were enriched in female mice and conserved in human recanalization models, pointing to potential sex-specific differences in reperfusion injury and stroke recovery mechanisms. The discovery of transcription factors NFE4 and MTF1, alongside miRNAs involved in reperfusion and recanalization injury, provides deeper insight into the regulatory dynamics of post-stroke recovery. These findings underscore the importance of targeting these molecular interactions in future therapeutic strategies. Experimental validation of these associations is essential, as it will advance understanding of stroke pathogenesis and potentially lead to novel, more effective treatments for ischemic stroke. By focusing on these molecular mechanisms, future research may enhance the efficacy of recanalization therapies, minimize futile recanalization, and ultimately improve patient outcomes in stroke recovery.

## Supporting information

Graphical Abstract

## Data Availability

Data will be made available after publication via GEO accession GSE278554: https://www.ncbi.nlm.nih.gov/geo/query/acc.cgi?acc=GSE278554

## Availability of data and materials

The dataset supporting the conclusions of this article is available in the GEO repository accession GSE278554.

## Author contributions

T.A.B., I.R.W, Y.M., and G.C.J designed experiments. T.A.B. and Y.M performed experiments and collected data, T.A.B. analyzed data. T.A.B., G.C.J. and I.R.W. wrote the main manuscript text, and T.A.B. prepared all the figures. All authors reviewed and revised the manuscript.

## Acknowledgements

The authors thank Sima Abbasi Habashi and Patricia Kent for their continual support.

## Ethics declaration

All experiments were approved by the University of Alberta’s Health Sciences Animal Care and Use Committee (Protocol AUP361) and adhered to the guidelines set by the Canadian Council for Animal Care.

## Supplemental Material

**Supplementary Table 1.** Differential gene expression and multivariate analysis results of MCAO, MCAO+Recan, Recan (MCAO+Recan vs. MCAO) (|log_2_FC| ≥1.5, p-value ≤ 0.05), Sex × MCAO+Recan interactions, and Age × MCAO+Recan interactions (|log_2_FC| ≥1.5, FDR ≤ 0.05). All analyses were performed using limma package in R Bioconductor.

**Supplementary Table 2.** Enrichment analysis gProfiler results for MCAO, MCAO+Recan, and Recan (MCAO+Recan vs. MCAO) groups. Columns include the group name, pathway category, term ID, description of the biological process, adjusted p-value (p.adjust), query size, count of genes in each pathway, total pathway size, effective domain size, and the list of associated genes (geneID). The gene ratio represents the proportion of query genes in the pathway, while the background ratio (BgRatio) represents the term size relative to the total background gene set.

**Supplementary Table 3.** The ARRIVE Guidelines 2.0: updated guidelines for reporting animal research. Form was obtained from https://www.goodreports.org/reporting-checklists/arrive2/ (28).

**Supplementary Figure 1.**
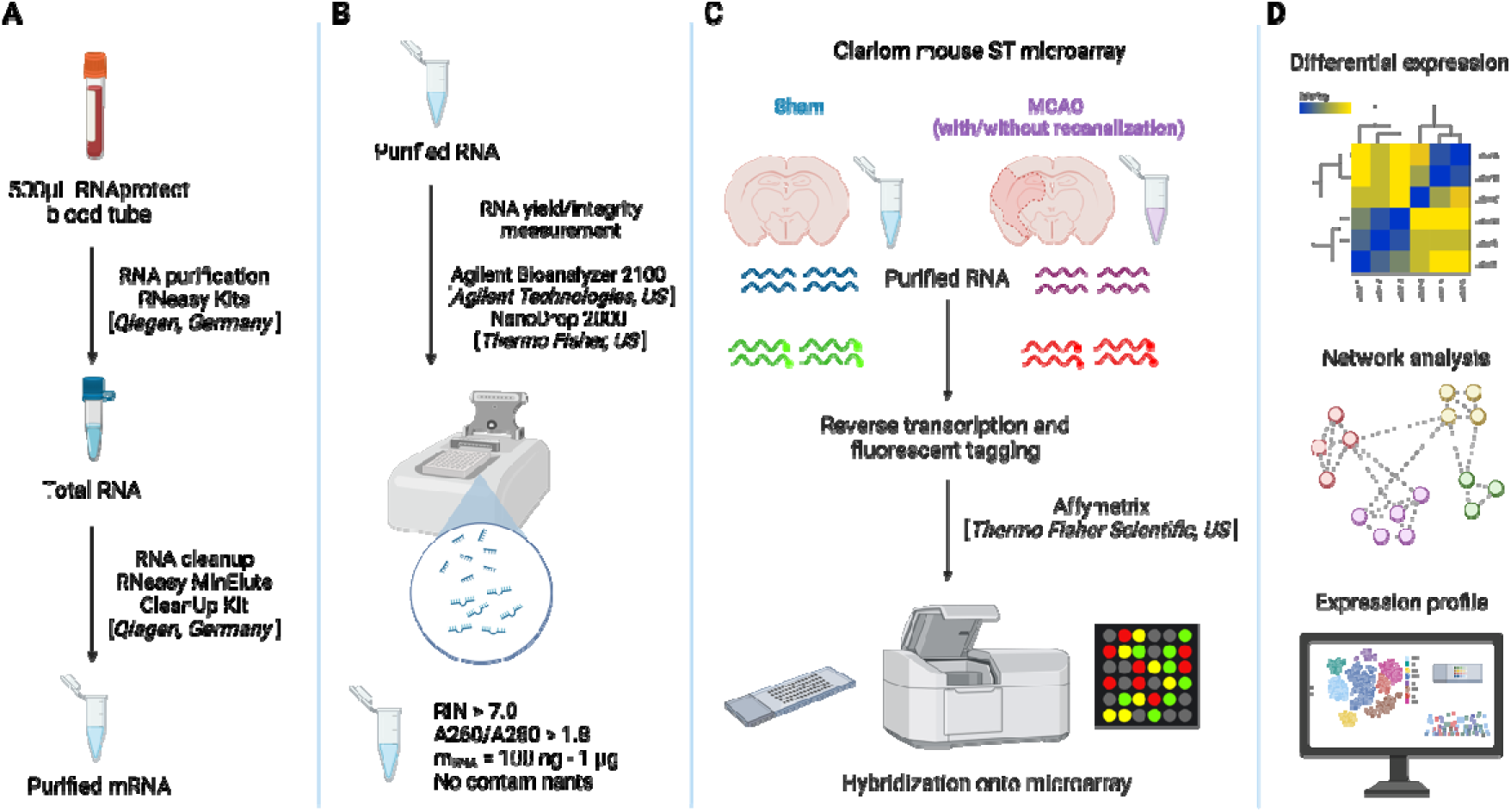
Overview of RNA purification and microarray analysis procedures. **(A)** Sample collection and RNA extraction. Whole blood samples were obtained via retro-orbital sampling following sham and MCAO surgeries. Total RNA was extracted and purified using standardized QIAGEN protocols to ensure high-quality RNA for downstream analyses. **(B)** RNA selection for quality control. RNA samples were rigorously evaluated for yield and purity to ensure that only high-quality samples were used for microarray analysis. Stringent quality control measures were applied to select samples with optimal RNA integrity. **(C)** Microarray analysis using Affymetrix technology. Selected RNA samples were processed and analyzed on Affymetrix microarrays to generate comprehensive gene expression profiles. **(D)** Data normalization and differential expression analysis. Raw CEL files generated from the Affymetrix platform were normalized using robust statistical methods, followed by differential gene expression analysis with the limma package in RStudio. Enrichment analysis was performed using the gProfiler tool to identify biologically significant pathways enriched among the differentially expressed genes. These enriched pathways were subsequently visualized and interpreted using RStudio and Cytoscape to explore the molecular mechanisms underlying the gene expression changes.

**Supplementary Figure 2.**
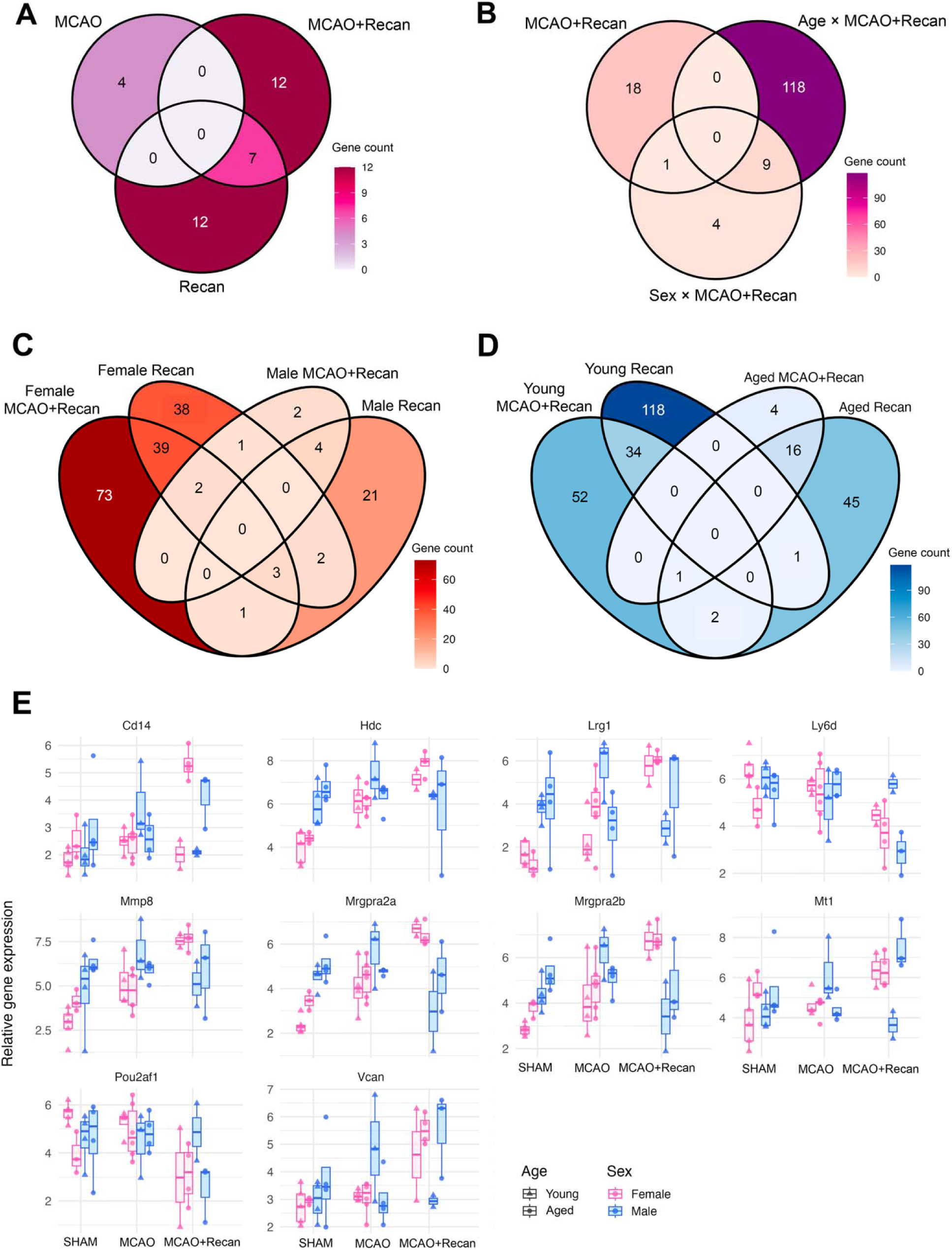
**(A)** Venn diagram showing the overlap of DEGs between MCAO, MCAO+Recan, and Recan groups (|log2FC| ≥ 1.5, p-value ≤ 0.05). **(B)** Venn diagram showing the overlap of DEGs between MCAO+Recan, Sex × Treatment and Age × Treatment interactions groups (|log2FC| ≥ 1.5, p-value ≤ 0.05). **(C)** Venn diagram showing the overlap of DEGs between sex-specific MCAO+Recan and Recan groups (|log2FC| ≥ 1.5, p-value ≤ 0.05). **(D)** Venn diagram showing the overlap of DEGs between age-specific MCAO+Recan and Recan groups (|log2FC| ≥ 1.5, p-value ≤ 0.05). All analyses were performed using limma package in R Bioconductor. **(E)** Sex- and age-specific expression patterns of differentially expressed genes in MCAO+Recan group: *Cd14, Mmp8, Hdc, Lrg1, Ly6d, Mrgpra2a, Mrgpra2b, Vcan, Mt1,* and *Pou2af1* determined by two-way interactions of Sex × Treatment and Age × Treatment multivariate analysis built-in in limma package (|log_2_FC| ≥ 1.5, p ≤ 0.05).

**Supplementary Figure 3.**
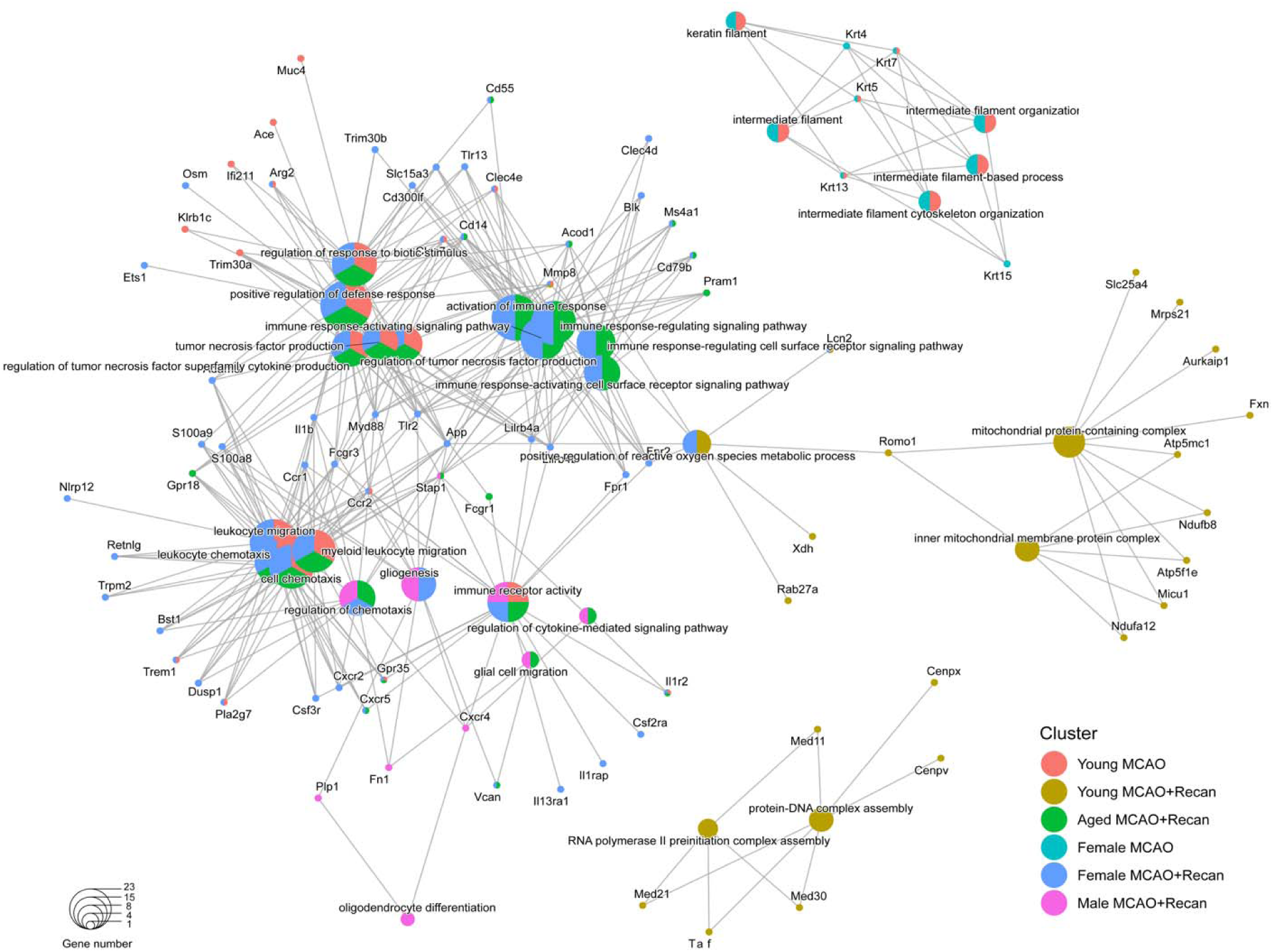
*cnet* plot showing gene overlaps in enriched gene sets. Enrichment analysis was performed using the *clusterProfiler* R package and *cnetplot()* function, with the Gene Ontology (GO) biological processes as the reference database. The plot visualizes the relationships between significantly enriched gene sets and their associated genes. Each node represents either a gene (small circles) or a gene set/pathway (larger colored circles). Edges indicate that a gene is part of a particular gene set. The size of each gene set node reflect the gene number.

**Supplementary Figure 4.**
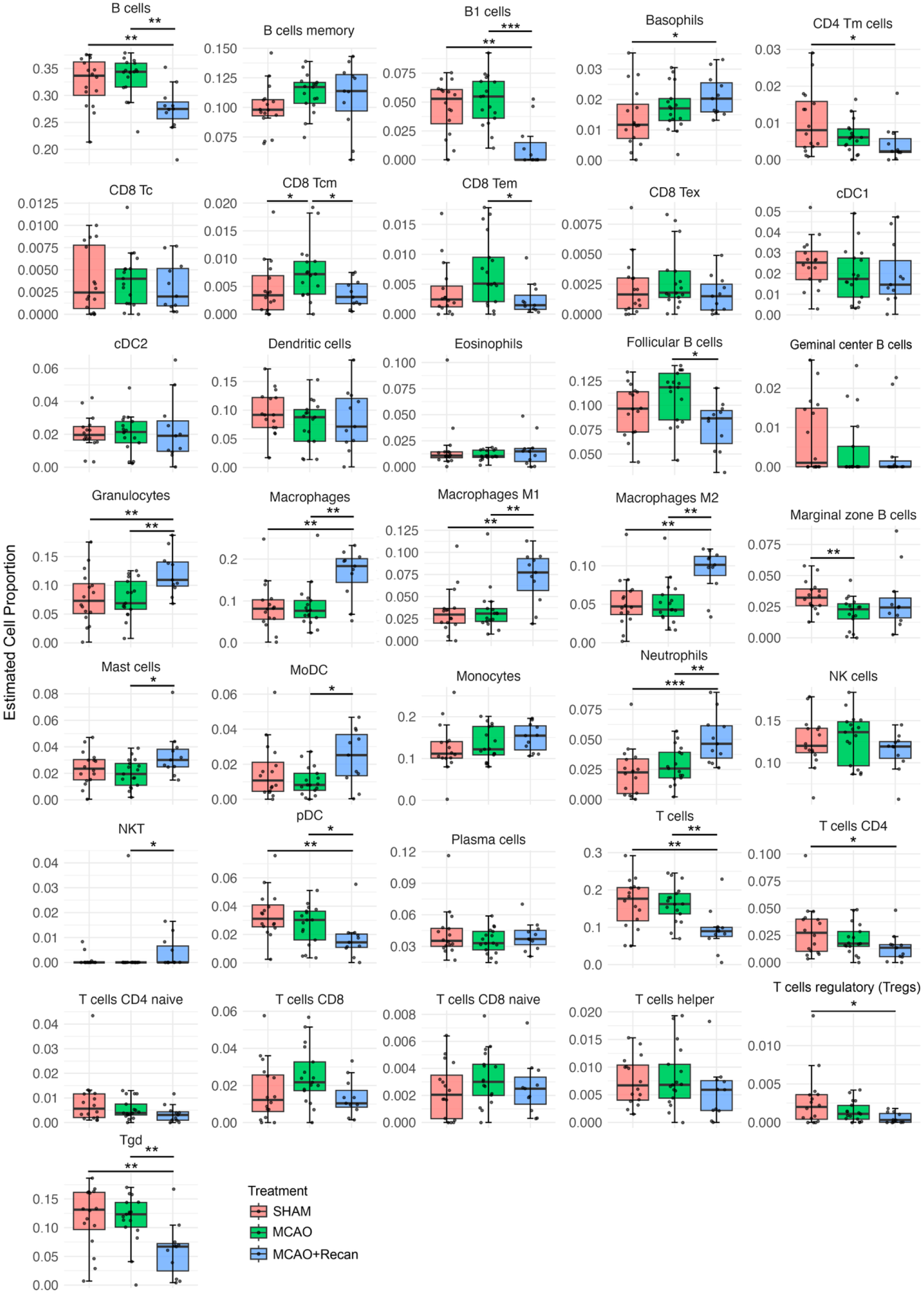
Relative immune cell proportions of 36 immune cell types estimated by ImmunCellAI-mouse, stratified by treatment group. Box plots display the estimated proportions of immune cell types. Statistical comparisons were performed using the Kruskal-Wallis, with p-values adjusted using the Benjamini-Hochberg method for multiple comparisons. Dunnett’s post-hoc pairwise multiple comparison was also performed (*p≤0.05, **p≤0.01, ***p≤0.001).

**Supplementary Figure 5.**
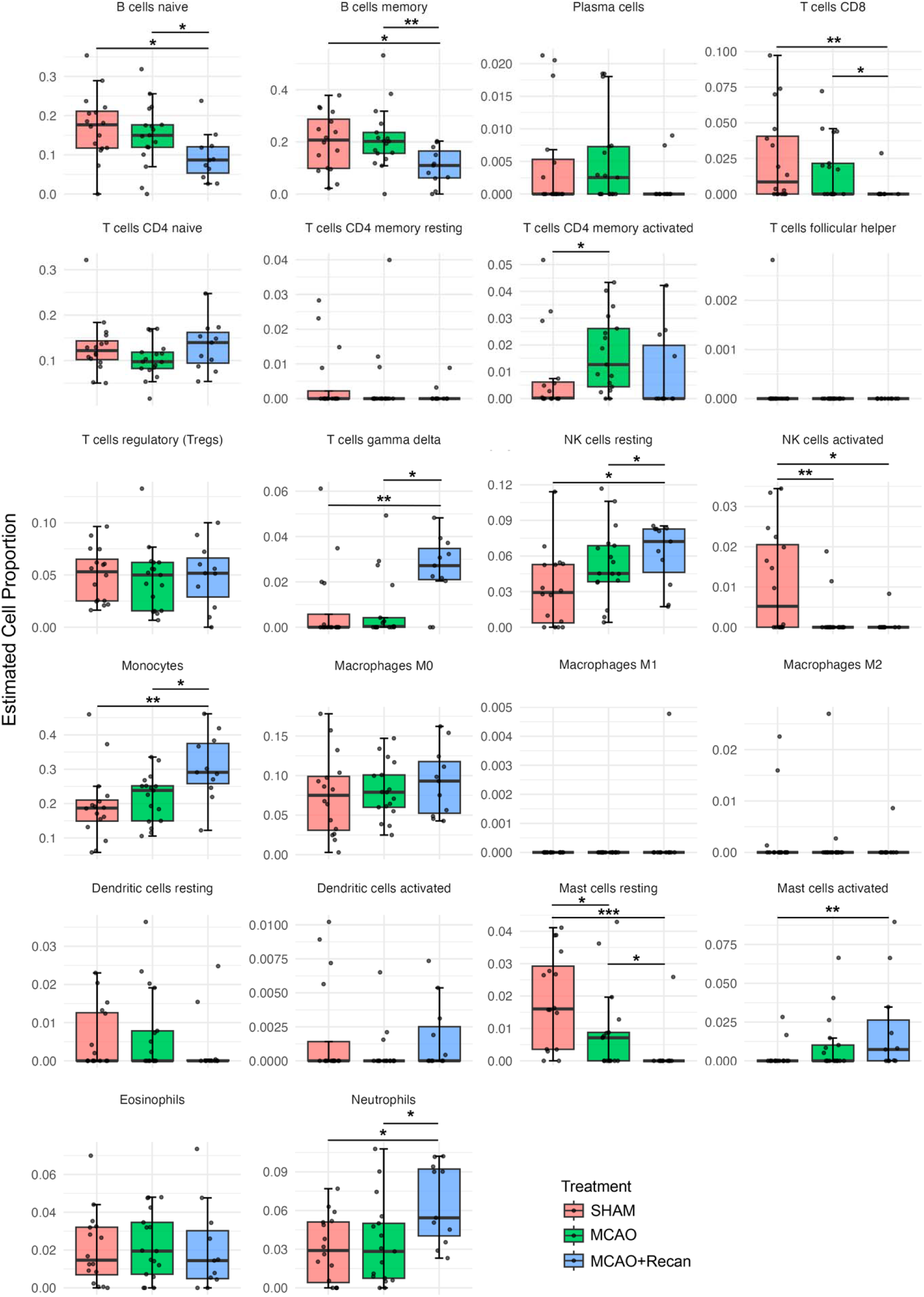
Relative immune cell proportions of 22 immune cells estimated by CIBERSORT, stratified by treatment group. Box plots display the estimated proportions of immune cell types inferred using the CIBERSORT algorithm based on bulk RNA-sequencing data and LM22 human reference signature matrix. Statistical comparisons were performed using the Kruskal-Wallis, with p-values adjusted using the Benjamini-Hochberg method for multiple comparisons. Dunnett’s post-hoc pairwise multiple comparison was also performed (*p≤0.05, **p≤0.01, ***p≤0.001).

**Supplementary Figure 6.**
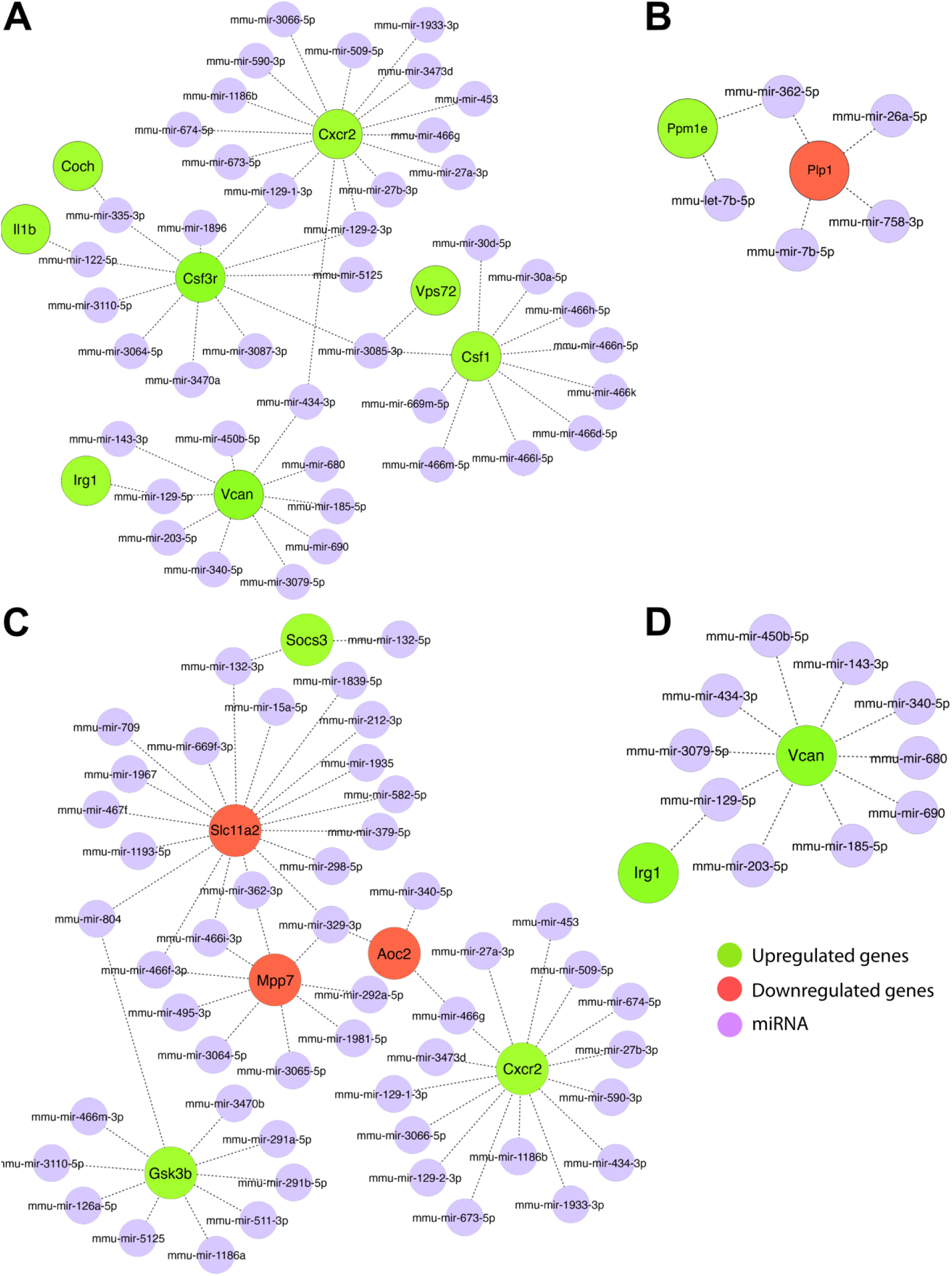
miRNA-mRNA interactions in **(A)** female, **(B)** male, **(C)** young, and aged MCAO+Recan animals from NetworkAnalyst generated from lists of DEGs. One gene may be regulated by multiple miRNAs, and each miRNA can target more than one gene.

**Supplementary Figure 7.**
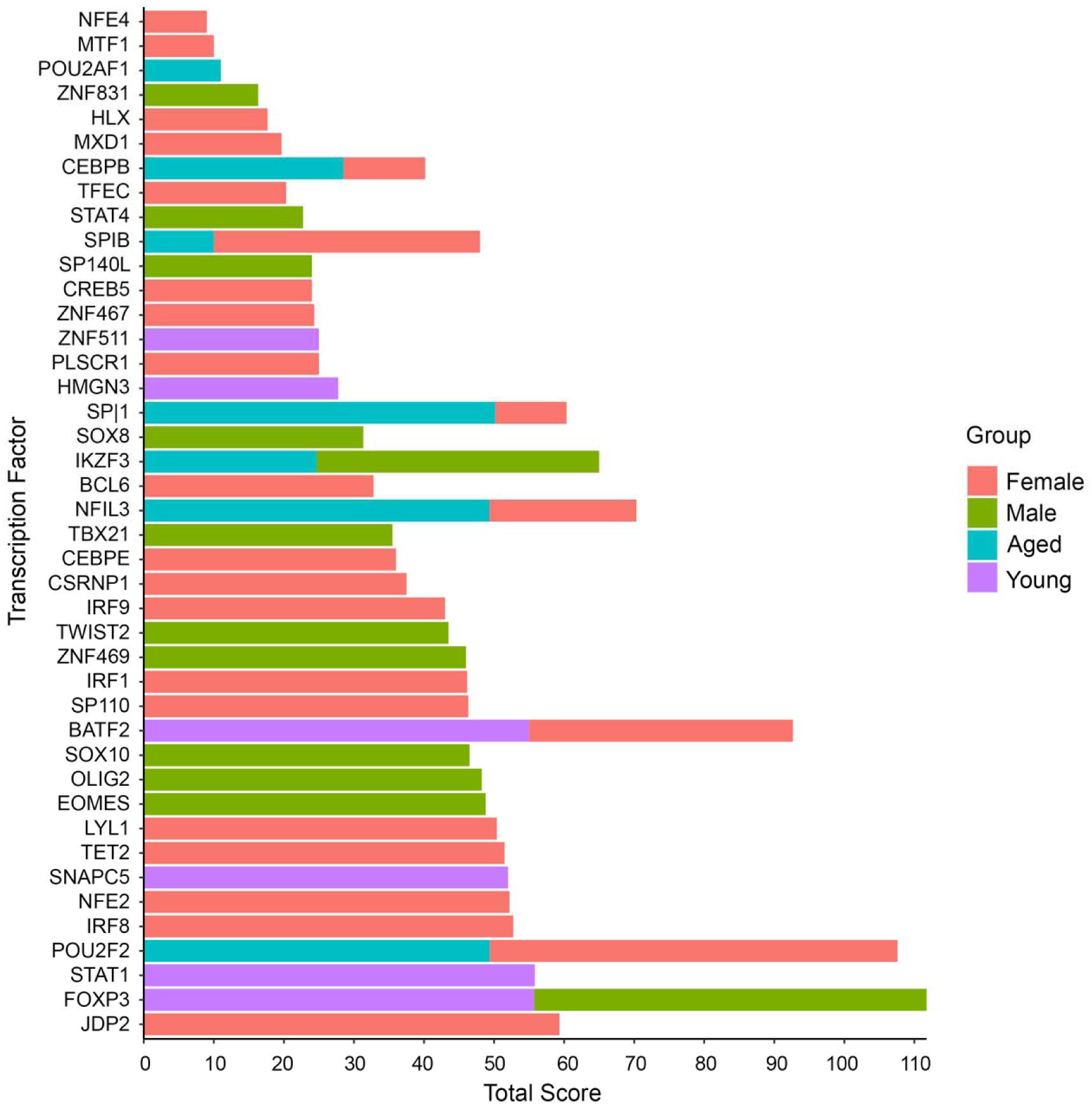
Top 50 TFs identified by ChEA3. ChEA3 was used to predict TFs-mRNA regulatory network. Lists of DEGs of MCAO+Recan were submitted to the ChEA3 platform, and the top 50 TFs (organized by rank) were exported. One TF can regulate several genes, and multiple TFs may regulate one gene.

