## Supplementary Table 3 for "Early transcriptional changes in neutrophil-mediated processes following recanalization after ischemic stroke"

### ARRIVE Reporting checklist for study using laboratory animals.

| | | Reporting Item | Page # |
|---|---|---|---|
| **Essential 10** | | | |
| Study design | #1a | Give details of the groups being compared, including control groups. If no control group has been used, the rationale should be stated. | 8-9 |
| Study design | #1b | Give details of the experimental unit (e.g., a single animal, litter, or cage of animals). | 8-9 |
| Sample size | #2a | Specify the exact number of experimental units allocated to each group, and the total number in each experiment. Also indicate the total number of animals used. | 8-9 |
| Sample size | #2b | Explain how the sample size was decided. Provide details of any a priori sample size calculation, if done. | 9, 27 |
| Inclusion and exclusion criteria | #3a | Describe any criteria used for including or excluding animals (or experimental units) during the experiment, and data points during the analysis. Specify if these criteria were established a priori. If no criteria were set, state this explicitly. | 9 |
| Inclusion and exclusion criteria | #3b | For each experimental group, report any animals, experimental units, or data points not included in the analysis and explain why. If there were no exclusions, state so. | 9 |
| Inclusion and exclusion criteria | #3c | For each analysis, report the exact value of n in each experimental group. | 8 |
| Randomisation | #4a | State whether randomisation was used to allocate experimental units to control and treatment groups. If done, provide the method used to generate the randomisation sequence. | 8-9 |
| Randomisation | #4b | Describe the strategy used to minimise potential confounders such as the order of treatments and measurements, or animal/cage location. If confounders were not controlled, state this explicitly. | 8-9 |
| Blinding | #5 | Describe who was aware of the group allocation at the different stages of the experiment (during the allocation, the conduct of the experiment, the outcome assessment, and the data analysis). | 9 |
| Outcome measures | #6a | Clearly define all outcome measures assessed (e.g., cell death, molecular markers, or behavioural changes). | 9-11 |
| Outcome measures | #6b | For hypothesis-testing studies, specify the primary outcome measure, i.e., the outcome measure that was used to determine the sample size. | 9-11 |
| Statistical methods | #7a | Provide details of the statistical methods used for each analysis, including software used. | 9-11 |
| Statistical methods | #7b | Describe any methods used to assess whether the data met the assumptions of the statistical approach, and what was done if the assumptions were not met. | 9-11 |
| Experimental animals | #8a | Provide species-appropriate details of the animals used, including species, strain and substrain, sex, age or developmental stage, and, if relevant, weight. | 8-9 |
| Experimental animals | #8b | Provide further relevant information on the provenance of animals, health/immune status, genetic modification status, genotype, and any previous procedures. | 8-9 |
| Experimental procedures | #9a | For each experimental group, including controls, describe the procedures in enough detail to allow others to replicate what was done, how it was done, and what was used. | 8-9 |
| Experimental procedures | #9b | Timing and frequency of procedures | 8-9 |

| | | | |
|---|---|---|---|
| Experimental procedures | #9c | Where procedures were carried out (including detail of any acclimatisation periods). | 8-9 |
| Experimental procedures | #9d | Rationale for procedures | 8-9 |
| Results | #10a | For each experiment conducted, including independent replications, report summary/descriptive statistics for each experimental group, with a measure of variability where applicable (e.g., mean and SD, or median and range). | 11-12 |
| Results | #10b | If applicable, for each experiment conducted, including independent replications, report the effect size with a confidence interval. | N/A |
| **Recommended set** | | | |
| Abstract | #11 | Provide an accurate summary of the research objectives, animal species, strain and sex, key methods, principal findings, and study conclusions. | 2 |
| Background | #12a | Include sufficient scientific background to understand the rationale and context for the study, and explain the experimental approach. | 5-8 |
| Background | #12b | Explain how the animal species and model used address the scientific objectives and, where appropriate, the relevance to human biology. | 8 |
| Objectives | #13 | Clearly describe the research question, research objectives and, where appropriate, specific hypotheses being tested. | 7-8 |
| Ethical statement | #14 | Provide the name of the ethical review committee or equivalent that has approved the use of animals in this study and any relevant licence or protocol numbers (if applicable). If ethical approval was not sought or granted, provide a justification. | 9 |
| Housing and husbandry | #15 | Provide details of housing and husbandry conditions, including any environmental enrichment. | 8 |
| Animal care and monitoring | #16a | Describe any interventions or steps taken in the experimental protocols to reduce pain, suffering, and distress. | 9 |
| Animal care and monitoring | #16b | Report any expected or unexpected adverse events. | N/A |
| Animal care and monitoring | #16c | Describe the humane endpoints established for the study, the signs that were monitored, and the frequency of monitoring. If the study did not set humane endpoints, state this. | 9 |
| Interpretation/scientific implications | #17a | Interpret the results, taking into account the study objectives and hypotheses, current theory, and other relevant studies in the literature. | 21-27 |
| Interpretation/scientific implications | #17b | Comment on the study limitations, including potential sources of bias, limitations of the animal model, and imprecision associated with the results. | 27-29 |
| Generalisability/translation | #18 | Comment on whether, and how, the findings of this study are likely to generalise to other species or experimental conditions, including any relevance to human biology (where appropriate). | 11, |
| Protocol registration | #19 | Provide a statement indicating whether a protocol (including the research question, key design features, and analysis plan) was prepared before the study, and if and where this protocol was registered. | N/A |
| Data access | #20 | Provide a statement describing if and where study data are available. | 30 |
| Declaration of interests | #21a | Declare any potential conflicts of interest, including financial and nonfinancial. If none exist, this should be stated. | 30 |
| Declaration of interests | #21b | List all funding sources (including grant identifier) and the role of the funder(s) in the design, analysis, and reporting of the study. | 30 |

None The ARRIVE checklist is distributed under the terms of the Creative Commons Attribution License CC-BY. This checklist can be completed online using https://www.goodreports.org/, a tool made by the EQUATOR Network in collaboration with Penelope.ai
