## Supplementary figures and images for "Early transcriptional changes in neutrophil-mediated processes following recanalization after ischemic stroke"

### Graphical Abstract

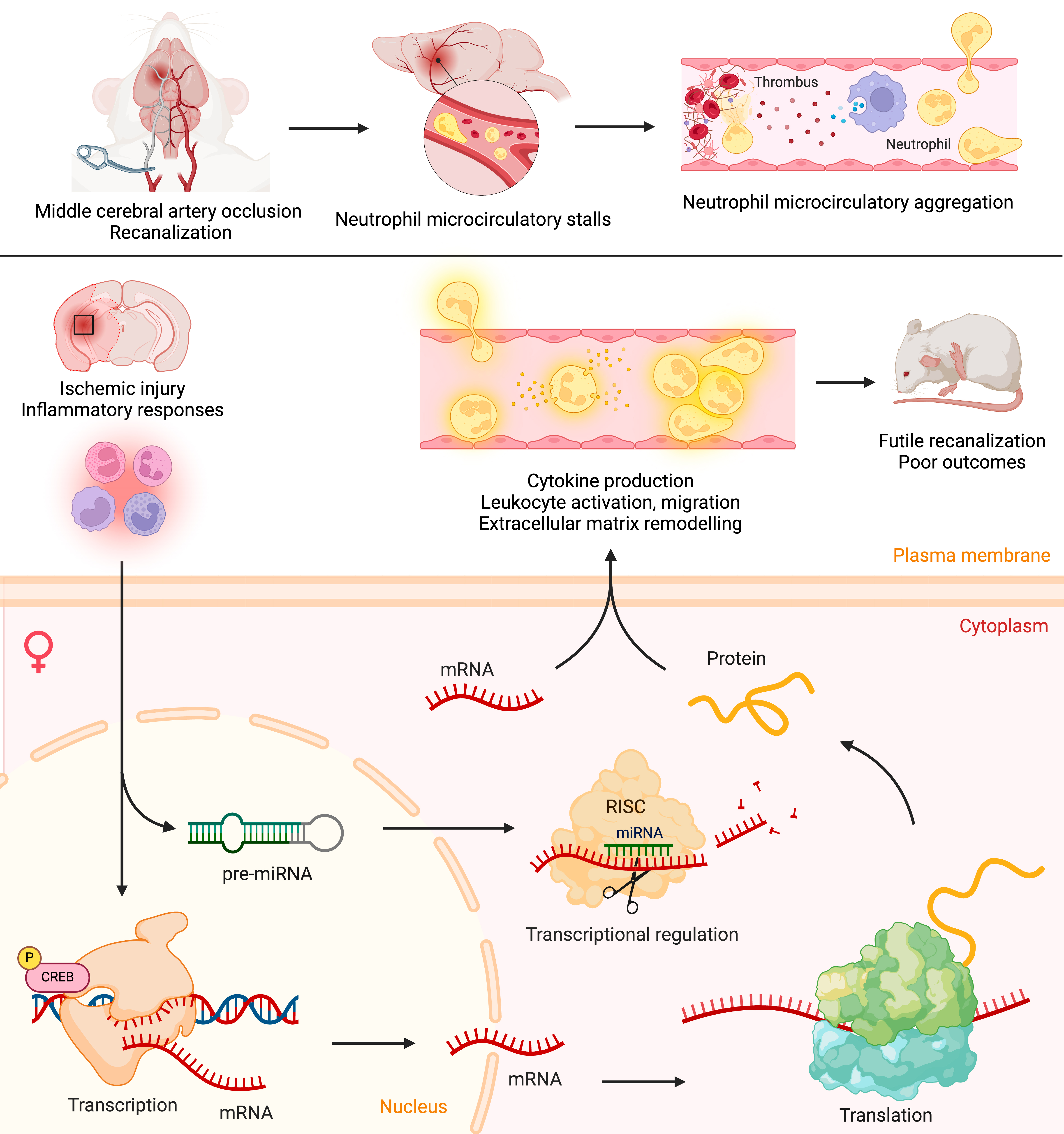

### Supplementary Figure 1

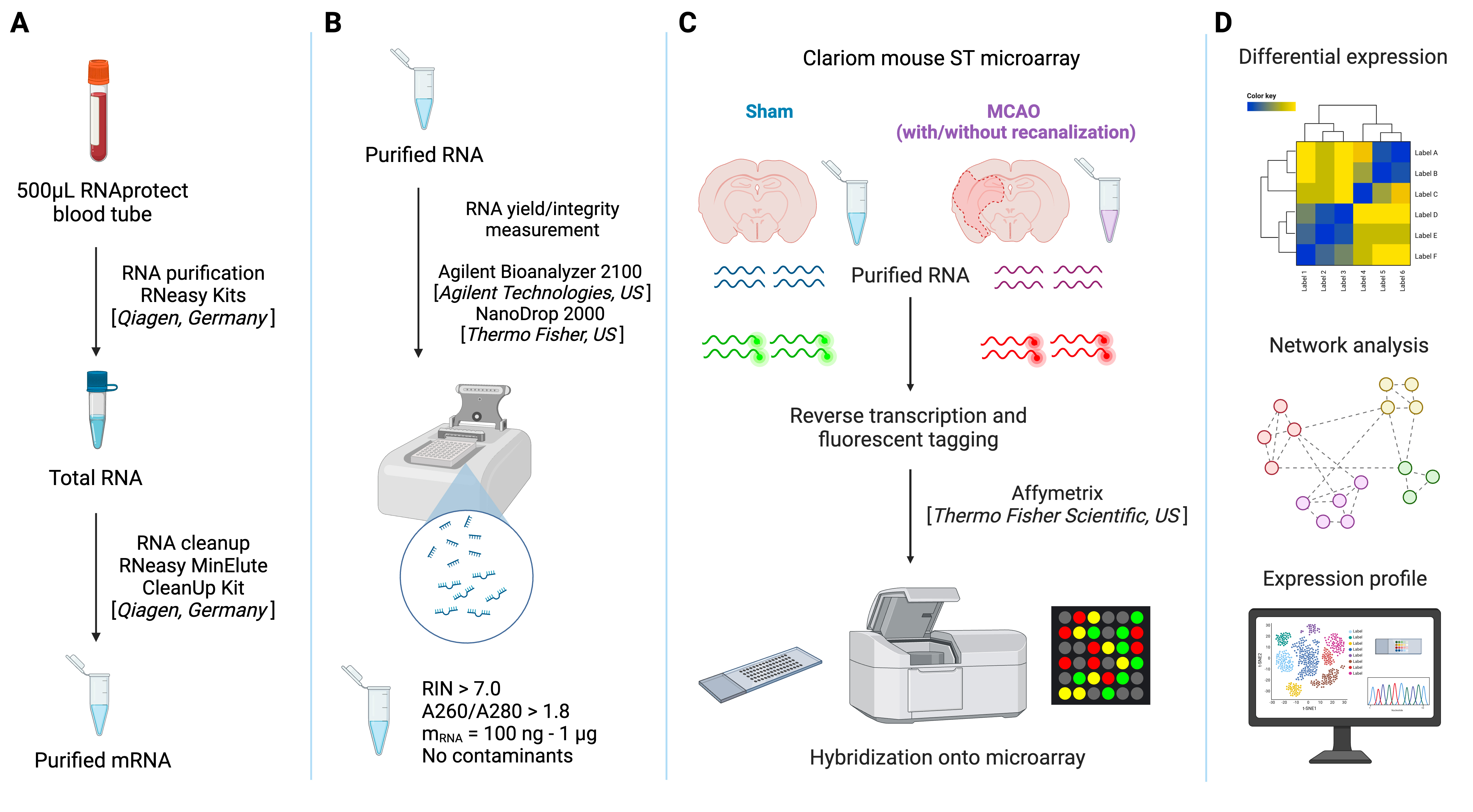

### Supplementary Figure 2

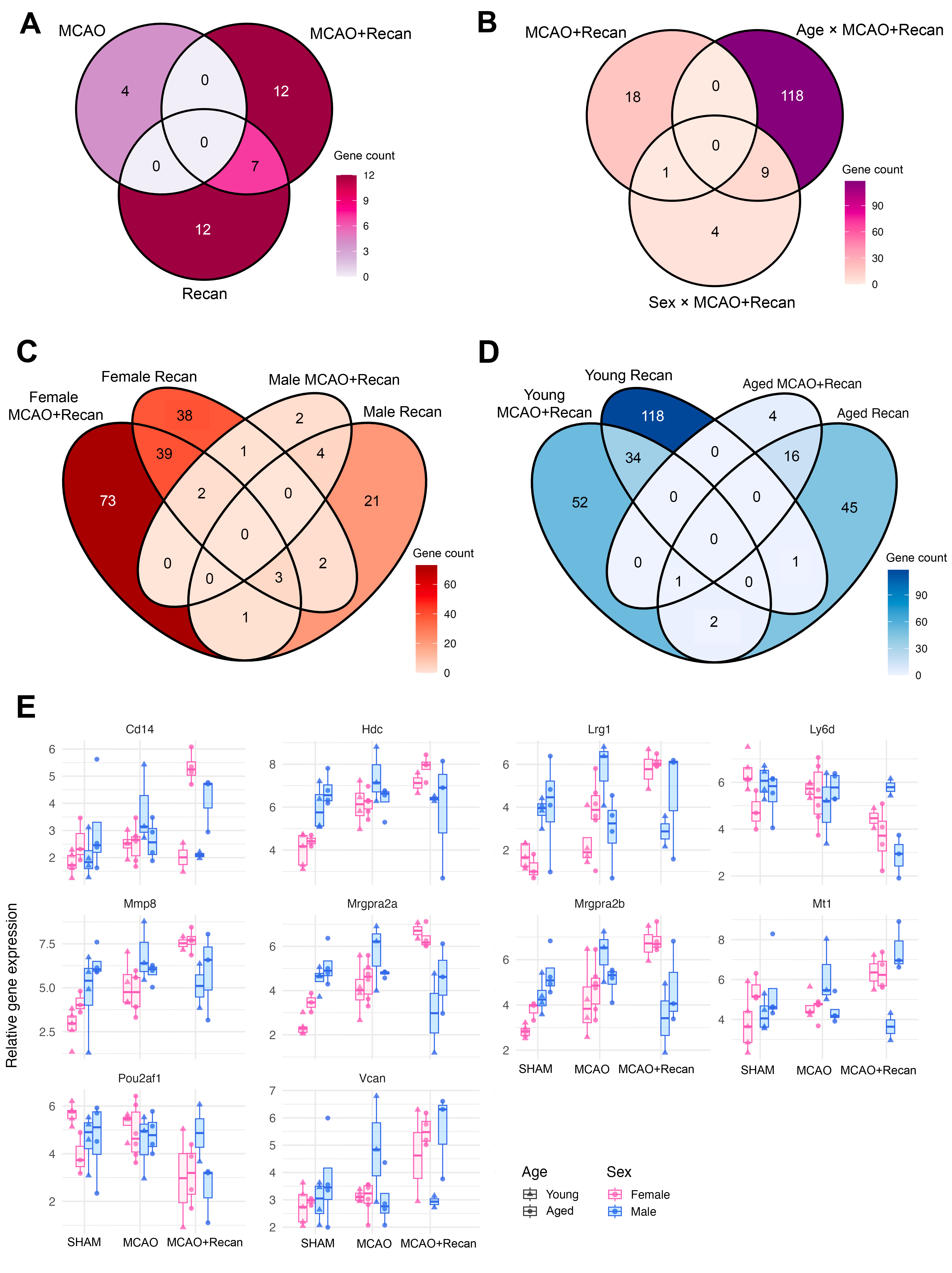

### Supplementary Figure 3

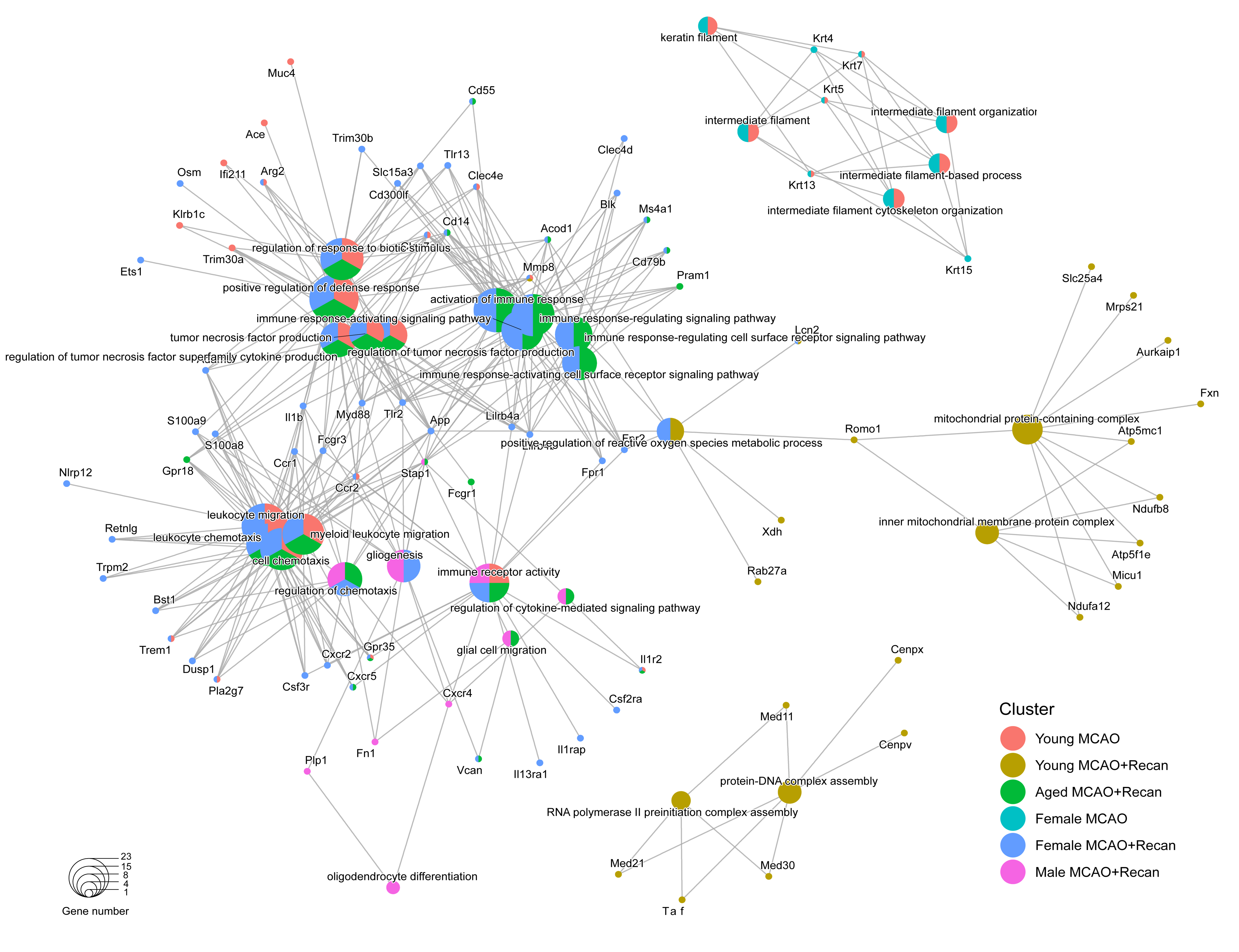

### Supplementary Figure 4

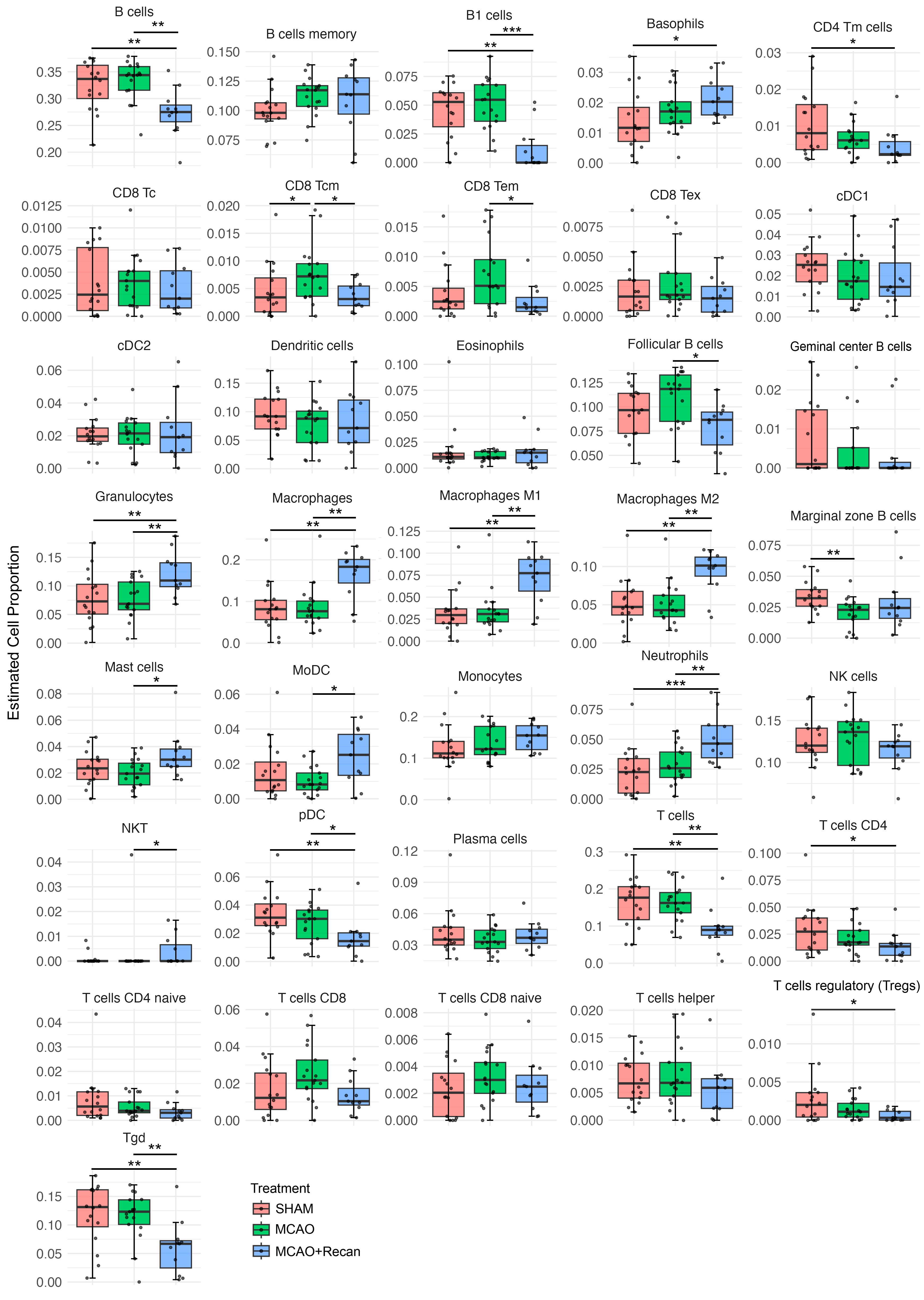

### Supplementary Figure 5

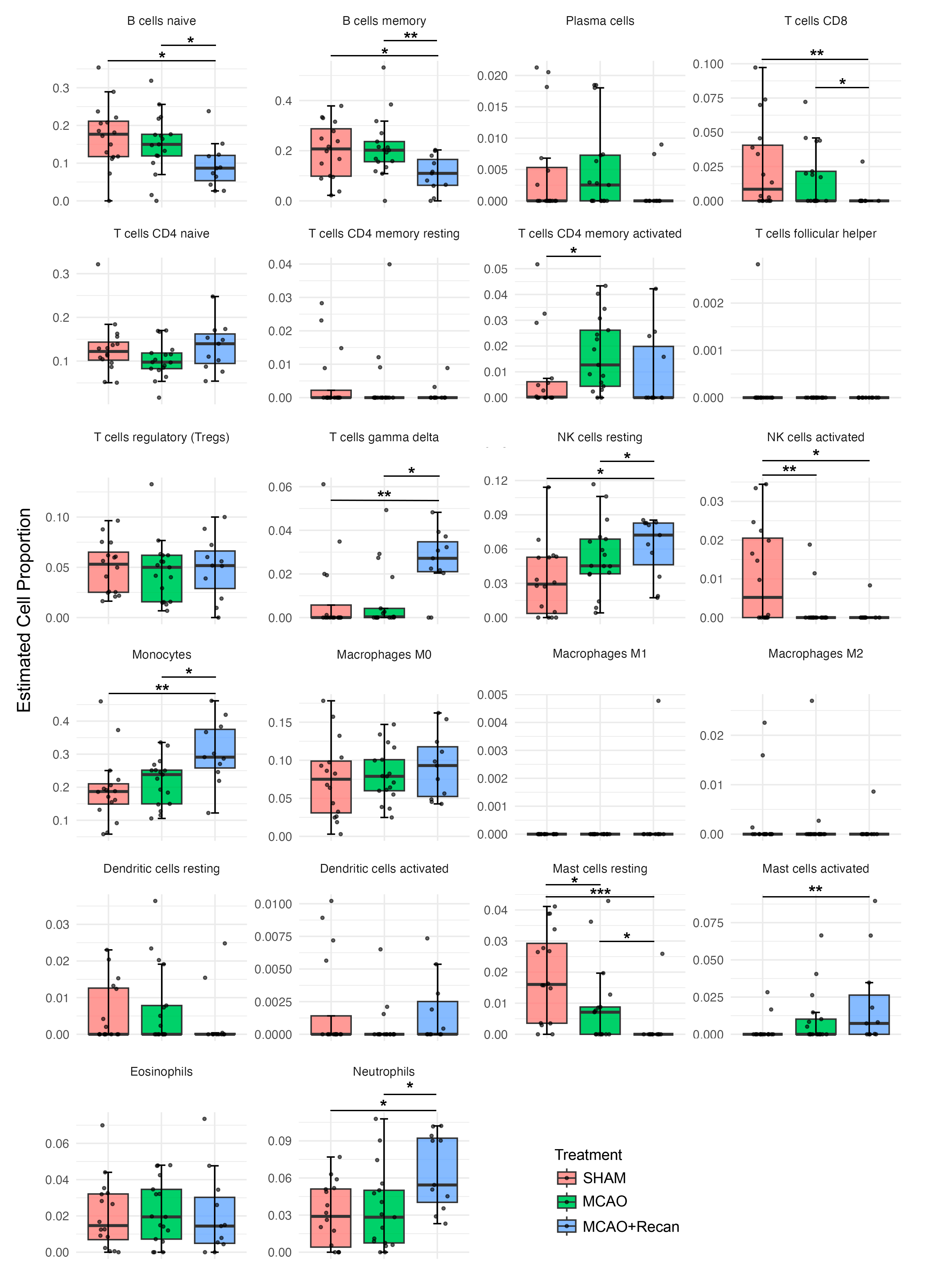

### Supplementary Figure 6

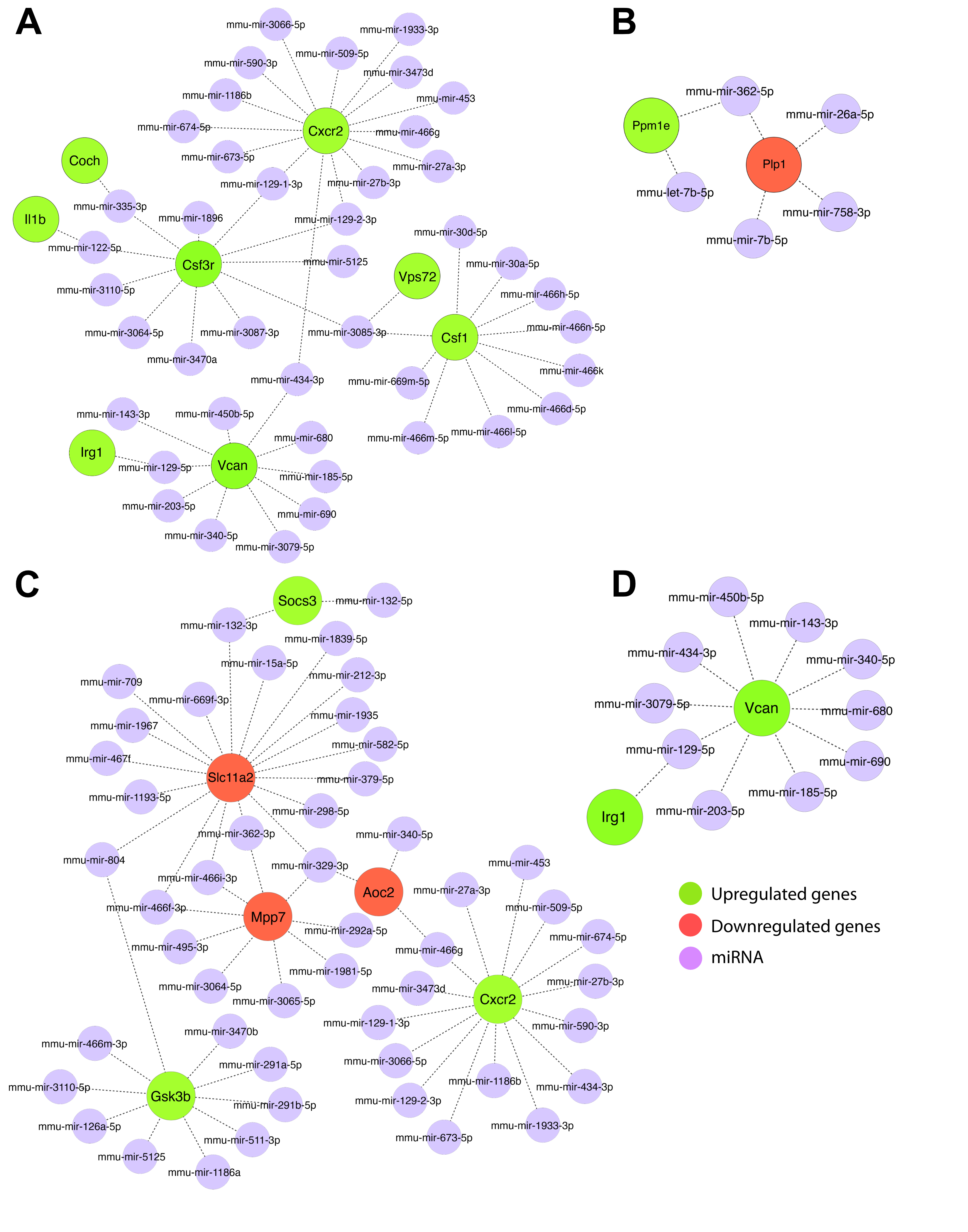

### Supplementary Figure 7

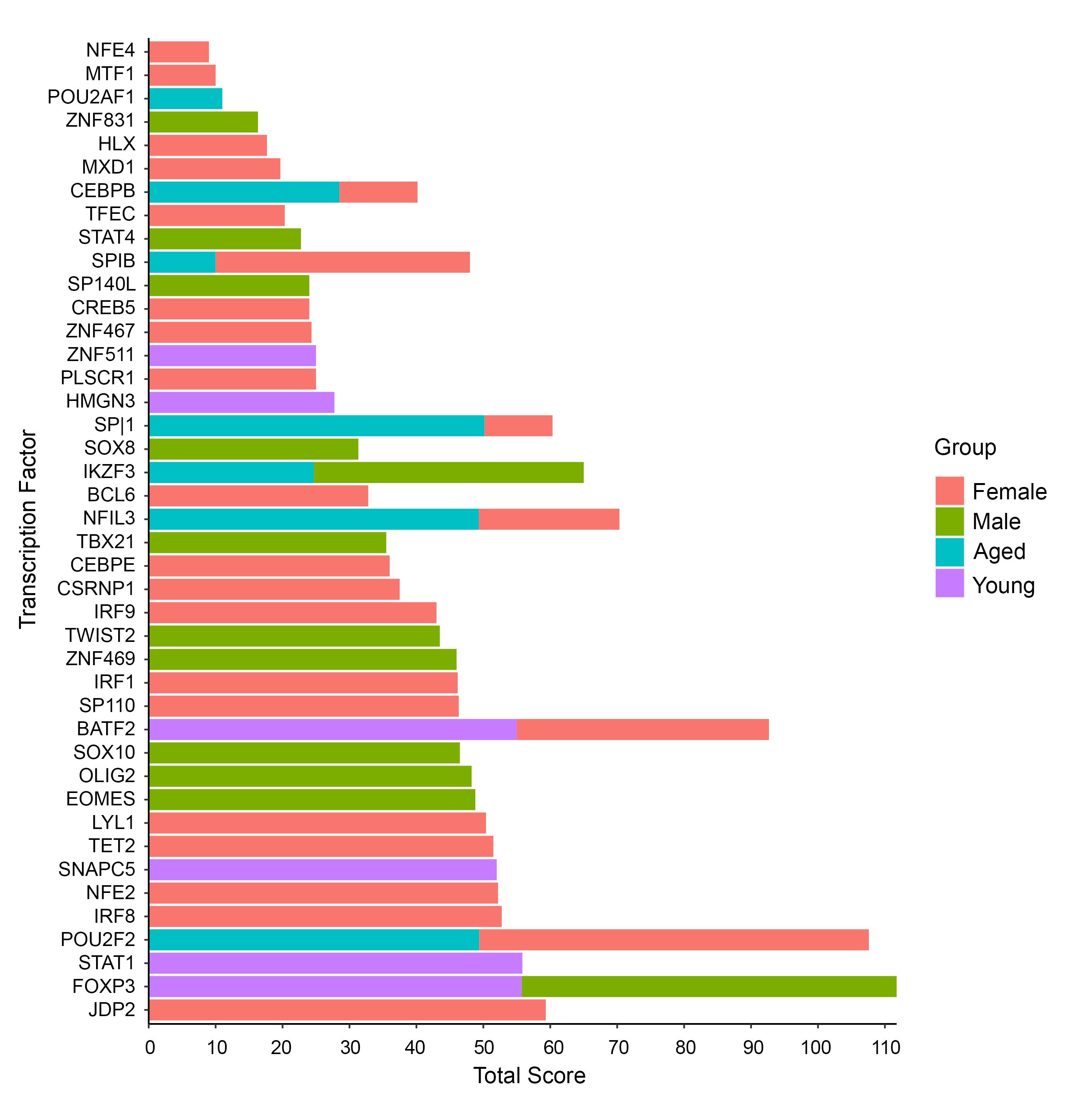
